# Causal associations of COVID-19 with health and disease outcomes: a systematic review and meta-analysis

**DOI:** 10.1101/2025.02.07.25321697

**Authors:** Lidong Gao, Junwei Yan, Mengfei Ye, Xiaopeng Yang, Yiying Yao, Jiajia Xia, Haonan Jin, Shanshan Ru, Mengdi Zhang, Zheng Liu, Yingzhe Zhang

## Abstract

**Objectives:** The COVID-19 pandemic has posed a substantial threat to global health. Despite numerous clinical observations and causality investigations, understanding of the disease’s progression and recovery process following infection remains limited. This systematic review and meta-analysis evaluates the causal connections between COVID-19 and various diseases using Mendelian randomization studies to provide robust theoretical underpinnings for the development of effective rehabilitation and intervention strategies.

**Methods:** We systematically searched PubMed, Embase, Web of Science, and Scopus for studies on Mendelian randomization related to COVID-19 published up to October 21, 2024. We used an extensive search strategy with the keywords “Mendelian randomization” and “COVID-19”. Two pairs of coauthors independently extracted data on study characteristics, exposure, and outcomes, resolving discrepancies through discussion. We included studies that considered susceptibility to COVID-19, hospitalization, or severe infection as the exposure and disease-related effects or impacts on human health as outcomes. We assessed the quality of the included studies using the MR-STROBE criteria and extracted the relative risk (odds ratio [OR]) using a random-effects model for meta-analysis. This study is registered with PROSPERO, CRD42025615426.

**Results:** Of the 1654 studies identified, 87 met the inclusion criteria for our meta-analysis. The primary outcome suggests that COVID-19 is associated with an increased risk of cardio-cerebral vascular diseases. Subgroup analyses identified an increased risk of neuropsychiatric disorders, including optic nerve disorders, epilepsy, schizophrenia, generalized anxiety disorder, stroke, myocardial infarction with COVID-19. There is also a notable association between COVID-19 and the immune system, particularly neuromyelitis optica spectrum disorders and myasthenia gravis.

**Conclusions:** This study demonstrates that COVID-19 infection has direct causal effects on human health or certain diseases at the genetic level, which may manifest as increased or decreased susceptibility to disease and changes in disease severity.

## Introduction

COVID-19 is a life-threatening infectious disease caused by SARS-CoV-2, first identified in Wuhan, China, in November 2019. Since then, it has spread rapidly around the world, leading the World Health Organization to classify it as a pandemic in March 2020.^1^ The clinical manifestations of COVID-19 range from asymptomatic infection to severe illness requiring intensive care, and occasionally resulting in death.^2^ Emerging evidence suggests that the clinical burden of COVID-19 extends beyond the acute phase, affecting patients’ quality of life and posing a significant challenge to global health.^3^

Our understanding of the COVID-19 pandemic has evolved with studies describing its epidemiology, clinical characteristics, pathophysiology, and complications. However, the long-term consequences of the disease remain largely unclear, despite many clinicians having observed a variety of extrapulmonary manifestations associated with COVID-19.^4^ A systematic review identified 84 clinical signs or persistent symptoms as most common among 45 studies of post-COVID-19 individuals, with newly emerged diseases such as cardiovascular dysfunction, urinary system symptoms, and neurological disorders occurring more frequently.^5^ Therefore, exploring the impact of COVID-19 on human physiology or disease progression has significant implications for patient prognosis and subsequent infections.

Numerous researchers have investigated the associations between COVID-19 and various systemic diseases or health outcomes, but a comprehensive synthesis of these studies is lacking. Conflicting findings emerge from studies examining the same outcomes. For instance, a bidirectional study suggests that the causal link between genetic predisposition to SARS-CoV-2 infection and cognitive decline might be mediated by systemic inflammation.^6^ Conversely, another study, employing a reverse Mendelian Randomization (MR) analysis of COVID-19, revealed a non-significant association between exposure and cognitive performance, suggesting a unidirectional relationship.^7^ These reports are inconclusive regarding both clinical observations and causal inference. However, We searched PubMed, Embase, Web of Science, and Scopus for studies relate to “COVID-19”, “meta-analysis” and “Mendelian randomization” up to October 21, 2024, no systematic review has evaluated the quality of this causal evidence, and previous meta-analyses have not included MR studies. To comprehensively assess and summarize the evidence for the causal role of COVID-19 in multiple diseases, we conducted a systematic review and meta-analysis of published MR studies.

## Methods

### Literature Search and Inclusion Criteria

We conducted a systematic search of PubMed, Scopus, Embase, and Web of science databases for studies related to COVID-19 and MR up to October 21, 2024. The search strategy incorporated the terms “Mendelian randomization” and “COVID-19”. We included original full-text articles that reported associations between COVID-19 and risks to various health outcomes, including the central nervous system disease, cardio-cerebrovascular system disease, digestive system disease, immune system disease, endocrine system disease, genitourinary system disease, tumors and infectious diseases. When multiple publications reported outcomes based on the same Genome-Wide Association Studies (GWAS), we selected the publication with the largest sample size for COVID-19. Studies that did not fall within the above systems were excluded due to the limited number of studies, which precluded meta-analysis their estimates with other eligible studies. The studies were performed in accordance with the PRIMA and STROBE-MR guidelines.

### Data Extraction and Quality Assessment

Data extraction was conducted for each study, capturing details such as the first author’s family name, publication year, the consortium or study providing genetic variants related to COVID-19 exposure, the consortium or study supplying genetic association estimates for the disease, sample size indicated by the number of cases and non-cases, and the relative risk estimate expressed as the the odds ratio (OR) with the corresponding 95% confidence interval (CI) for the association with COVID-19. These estimates were derived from the main analysis using the inverse-variance weighted method, sensitivity analyses using the weighted median and MR-Egger methods, and a multivariable MR analysis adjusted for genetically predicted COVID-19. Data extraction was performed by one investigator and verified by another.

To ensure the transparency and quality of the research, we referred to the STROBE-MR guidelines for assessing the quality of included MR studies.

### Data Analysis

For outcomes with multiple MR estimates from non-overlapping samples, we derived a pooled estimate through meta-analysis using the metan command in Stata (version 12). We employed a random-effects model to estimate the impact of COVID-19 on human health or the disease progression. Most reported estimates were expressed in terms of the per standard deviation increase in the risk of COVID-19 susceptibility, hospitalization, or severe infection. Heterogeneity across studies was assessed using the I² statistic. We interpreted I² values of 25-50% as indicating mild heterogeneity, 50-75% as moderate heterogeneity, and >75% as severe heterogeneity. Publication bias was evaluated through visual inspection of the forest plots, and statistical significance was set at a two-tailed P-value of less than 0.05 for all analyses. For outcomes with considerable heterogeneity (I² between 50% and 75%), subgroup analysis was conducted to explore tween-study heterogeneity. Sensitivity analysis included outlier detection and influence diagnostics, such as leave-one-out cross-validation.

## Results

### Literature search, study selection

Our database search yielded 1654 records, which were reduced to 516 after removing duplicates. After excluding studies with overlapping or identical outcome data, 87 articles based on non-overlapping populations were eligible for inclusion in the meta-analyses (Figure 1). The distribution of included studies across outcome categories was as follows: 23 for central nervous system disease,^7–30^ 16 for cardio-cerebrovascular system,^31–46^ 5 for digestive system disease,^47–52^ 15 for autoimmune disease,^53–67^ 2 for endocrine system disease,^68 69^ 8 for genitourinary system disease,^70–77^ 12 for tumors,^78–89^ 3 for infectious diseases,^90–92^ 3 for multisystem disease.^93–95^ The quality assessment and results from the studies included in this review are shown in eTable 2. The methodological quality of the studies included was assessed as moderate.

**Fig. 1.**
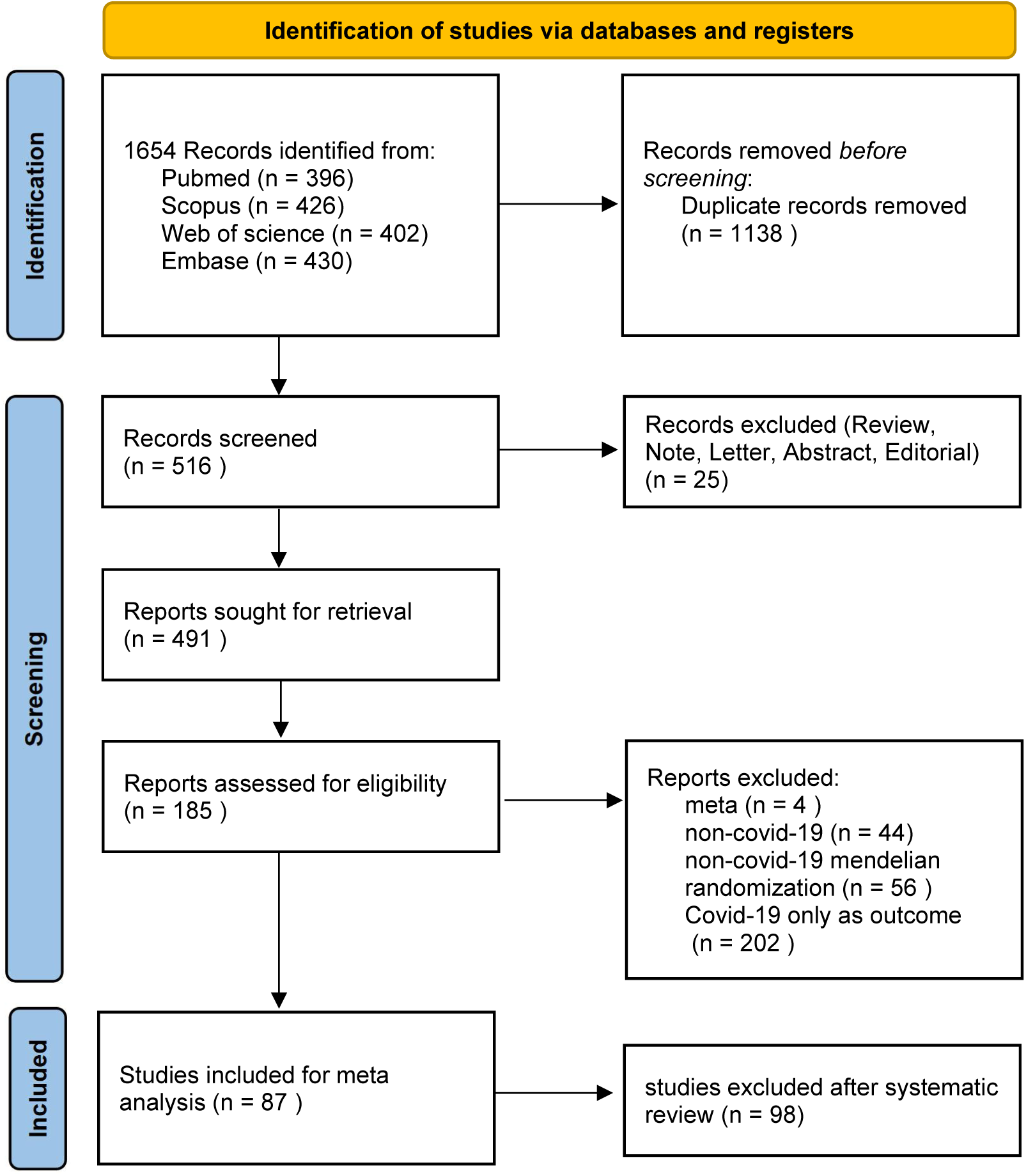
Flowchart of literature search, MR and meta-analysis. This flowchart provides a detailed overview of the research process. It includes the processes of literature search, screening, eligibility assessment, and the number of data points included in the meta-analysis. Source: Page MJ, et al. BMJ 2021;372:n71. doi: 10.1136/bmj.n71. This work is licensed under CC BY 4.0. To view a copy of this license, visit https://creativecommons.org/licenses/by/4.0/

### Study Description

The characteristics of the MR studies indicating causal relationships are presented in eTable 1. Out of 87 studies, 18 employed a bidirectional MR study design, allowing for the examination of causality in both directions. In most studies, the genetic variations of exposure originated from the COVID-19 Host Genetics Initiative. Although the version of the data and, consequently, the sample size varied across studies, this variation did not impact the overall findings.

### Meta-Analysis Results

Mendelian randomized studies^31–46 95^ evaluating the effects of COVID-19 on the cardio-cerebrovascular system showed a positive correlation between genetic predisposition and the risk of COVID-19 (OR 1.013, 95% CI 1.006-1.019). Furthermore, the risk of COVID-19 was positively correlated with pre-existing cardiovascular and cerebrovascular disease (OR 1.019, 95% CI 1.011-1.027). (Figure 2)

**Fig. 2.**
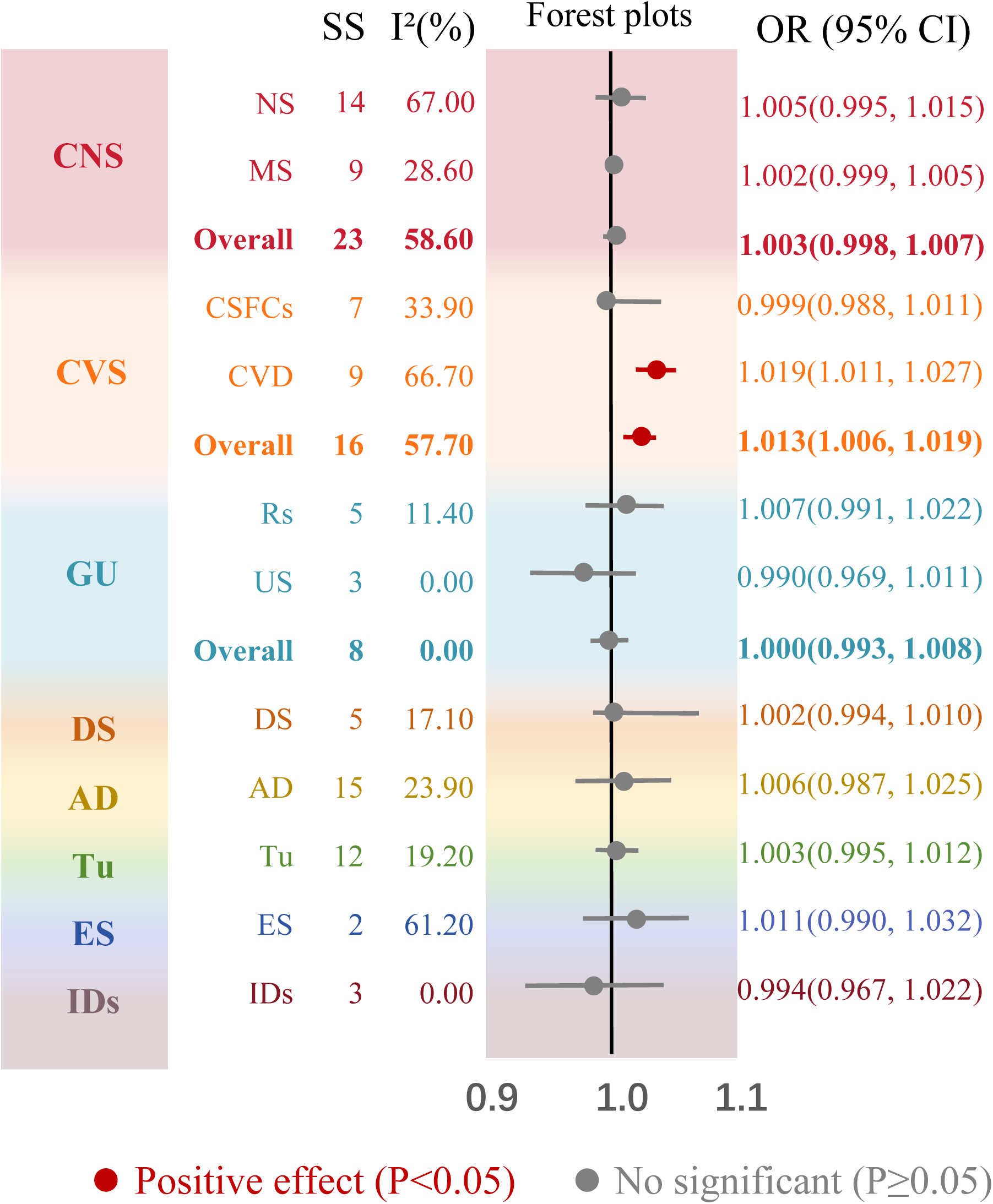
Forest plot of meta-analysis on the impact of COVID-19 on major human systems. AD, Autoimmunity diseases; CI, Confidence intervals; CNS, Central nervous system; CSFCs, Cardiac structural functional changes; CVD, Cardiovascular diseases; CVS, Cardiovascular and cerebrovascular diseases; DS, Digestive system; ES, Endocrinium diseases; GU, Genitourinary diseases; IDs, Infectious diseases; MS, Mental system diseases; NS, Nervous system diseases; OR, Odds ratio; Rs, Reproductive system; Tu, Tumour; US, Urinary system.

### Subgroup analysis

We identified significant heterogeneity in some of the meta-analysis results. To determine the source of this variability, we performed subgroup analyses to further explore the causal relationship between COVID-19 and the disorders (eTable 3).

In the subgroup analyses (Figure 3), a positive correlation was observed between COVID-19 and epilepsy, with an OR of 1.087 and a 95% CI ranging from 1.053 to 1.122.^16 20 22 23 28^ We also analyzed the three severity levels of COVID-19 with epilepsy (Table 1), finding that susceptibility to COVID-19, hospitalization status, and severe infection had a positive impact on epilepsy, with ORs of 1.081 (95% CI 1.021-1.167), 1.092 (95% CI 1.033-1.151) and 1.066 (95% CI 1.015-1.116), respectively.

**Fig. 3.**
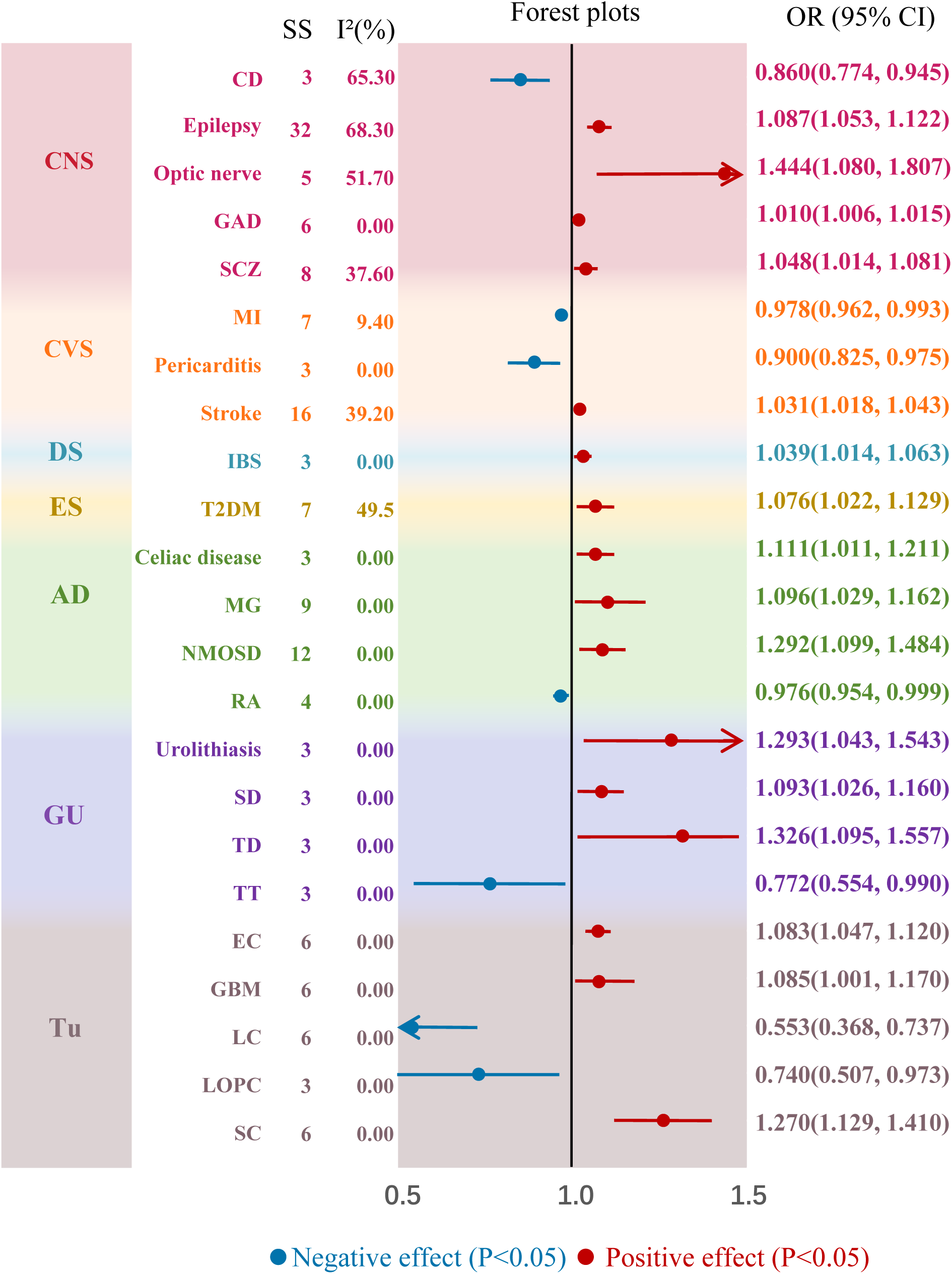
Subgroup analysis results. This figure, which includes a heatmap and a forest plot, reflects the subgroup results of a MR meta-analysis on the effects of three B vitamins on fifteen neuropsychiatric disorders. AD, Autoimmunity dieases; CI, Confidence intervals; CNS, Central nervous system; CVS, Cardiovascular and cerebrovascular diseases; DS, Digestive system; EC, Endometrial cancer; ES, Endocrinium dieases; GAD, Generalized anxiety disorder; GBM, Glioblastoma multiforme; GU, Genitourinary dieases; IBS, Irritable bowel syndrome; MG, Myasthenia gravis; MI, Myocardial infarction; NMOSD, Neuromyelitis optica spectrum disorder; OR, Odds ratio; RA, Rheumatoid Arthritis; SCZ, Schizophrenia; Tu, Tumor; T2DM, Type 2 Diabetes mellitus.

**Table 1.**
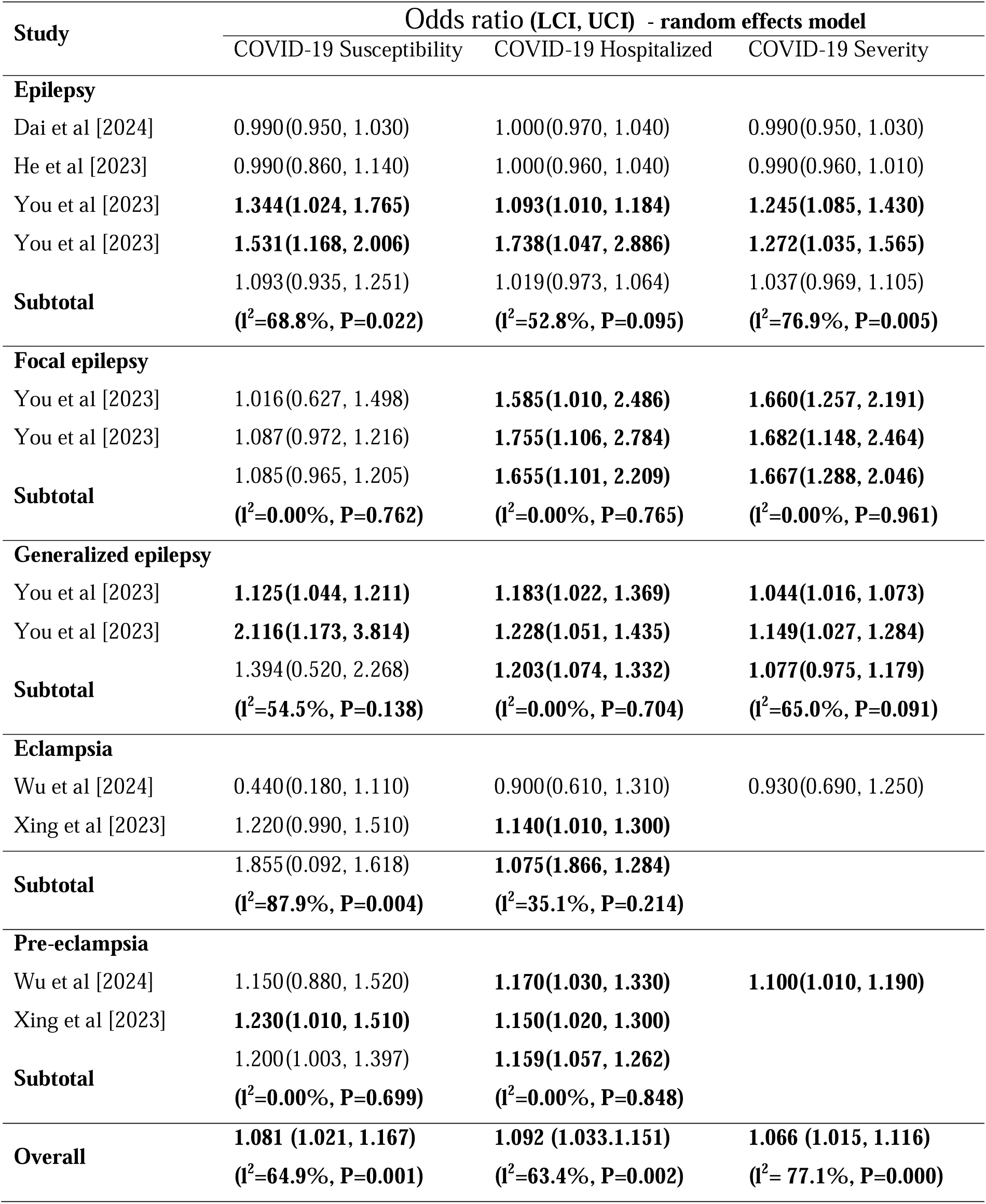
Positive causal relationship between COVID-19 and epilepsy through subgroup meta-analysis.

| Study | Odds ratio (LCI, UCI) - random effects model |  |  |
| --- | --- | --- | --- |
|  | COVID-19 Susceptibility | COVID-19 Hospitalized | COVID-19 Severity |
| **Epilepsy** |  |  |  |
| Dai et al [2024] | 0.990(0.950, 1.030) | 1.000(0.970, 1.040) | 0.990(0.950, 1.030) |
| He et al [2023] | 0.990(0.860, 1.140) | 1.000(0.960, 1.040) | 0.990(0.960, 1.010) |
| You et al [2023] | **1.344(1.024, 1.765)** | **1.093(1.010, 1.184)** | **1.245(1.085, 1.430)** |
| You et al [2023] | **1.531(1.168, 2.006)** | **1.738(1.047, 2.886)** | **1.272(1.035, 1.565)** |
| **Subtotal** | 1.093(0.935, 1.251)<br>**(I<sup>2</sup>=68.8%, P=0.022)** | 1.019(0.973, 1.064)<br>**(I<sup>2</sup>=52.8%, P=0.095)** | 1.037(0.969, 1.105)<br>**(I<sup>2</sup>=76.9%, P=0.005)** |
| **Focal epilepsy** |  |  |  |
| You et al [2023] | 1.016(0.627, 1.498) | **1.585(1.010, 2.486)** | **1.660(1.257, 2.191)** |
| You et al [2023] | 1.087(0.972, 1.216) | **1.755(1.106, 2.784)** | **1.682(1.148, 2.464)** |
| **Subtotal** | 1.085(0.965, 1.205)<br>**(I<sup>2</sup>=0.00%, P=0.762)** | 1.655(1.101, 2.209)<br>**(I<sup>2</sup>=0.00%, P=0.765)** | 1.667(1.288, 2.046)<br>**(I<sup>2</sup>=0.00%, P=0.961)** |
| **Generalized epilepsy** |  |  |  |
| You et al [2023] | **1.125(1.044, 1.211)** | **1.183(1.022, 1.369)** | **1.044(1.016, 1.073)** |
| You et al [2023] | **2.116(1.173, 3.814)** | **1.228(1.051, 1.435)** | **1.149(1.027, 1.284)** |
| **Subtotal** | 1.394(0.520, 2.268)<br>**(I<sup>2</sup>=54.5%, P=0.138)** | 1.203(1.074, 1.332)<br>**(I<sup>2</sup>=0.00%, P=0.704)** | 1.077(0.975, 1.179)<br>**(I<sup>2</sup>=65.0%, P=0.091)** |
| **Eclampsia** |  |  |  |
| Wu et al [2024] | 0.440(0.180, 1.110) | 0.900(0.610, 1.310) | 0.930(0.690, 1.250) |
| Xing et al [2023] | 1.220(0.990, 1.510) | **1.140(1.010, 1.300)** |  |
| **Subtotal** | 1.855(0.092, 1.618)<br>**(I<sup>2</sup>=87.9%, P=0.004)** | 1.075(1.866, 1.284)<br>**(I<sup>2</sup>=35.1%, P=0.214)** |  |
| **Pre-eclampsia** |  |  |  |
| Wu et al [2024] | 1.150(0.880, 1.520) | **1.170(1.030, 1.330)** | **1.100(1.010, 1.190)** |
| Xing et al [2023] | **1.230(1.010, 1.510)** | **1.150(1.020, 1.300)** |  |
| **Subtotal** | 1.200(1.003, 1.397)<br>**(I<sup>2</sup>=0.00%, P=0.699)** | 1.159(1.057, 1.262)<br>**(I<sup>2</sup>=0.00%, P=0.848)** |  |
| **Overall** | **1.081 (1.021, 1.167)**<br>**(I<sup>2</sup>=64.9%, P=0.001)** | **1.092 (1.033,1.151)**<br>**(I<sup>2</sup>=63.4%, P=0.002)** | **1.066 (1.015, 1.116)**<br>**(I<sup>2</sup>= 77.1%, P=0.000)** |

Interestingly, hospitalization and severe COVID-19 infection were positively associated with both focal and generalized epilepsy, with ORs of 1.655 (95% CI 1.101-2.209), 1.667 (95% CI 1.288-2.046), 1.203 (95% CI 1.074-1.332) and 1.077 (95% CI 0.975-1.179), respectively.

Additionally, hospitalization due to COVID-19 showed a positive effect on eclampsia and pre-eclampsia, with ORs of 1.075 (95% CI 1.866-1.284) and 1.159 (95% CI 1.057-1.262), respectively.

Conversely, COVID-19 demonstrated a negative correlation with cognitive dysfunction^96^ with ORs of 0.86 (95% CI 0.774-0.945). Furthermore, COVID-19 was found to be positively correlated with optic nerve disorders, schizophrenia and generalized anxiety disorder, with ORs of 1.444 (95% CI 1.080-1.807),^25 30^ 1.048 (95% CI 1.014-1.081)^8 9 21^ and 1.01 (95% CI 1.006-1.015)^9 21^, respectively (Figure 3).

In the context of the cardiovascular and cerebrovascular systems, COVID-19 has been found to have a positive association with stroke. The OR for stroke was 1.031 with a 95% CI of 1.018 to 1.043.^23 32 37 40^ In contrast, COVID-19 exhibited a negative correlation with myocardial infarction and pericarditis, both with ORs of 0.978 and a 95% CI of 0.962 to 0.993 and 0.900 and a 95% CI of 0.825 to 0.975 (Figure 3).

In the digestive system, the meta-analysis outcomes indicated a positive correlation between COVID-19 and irritable bowel syndrome, with an OR of 1.039 and a 95% CI of 1.014-1.063.^50^ Additionally, COVID-19 was found to have a positive correlation with type 2 diabetes mellitus, with an OR of 1.076 and a 95% CI of 1.022-1.129.^69 95^ (Figure 3)

While COVID-19 has no overall effect on the immune system, our meta-analysis results indicated that COVID-19 was positively correlated with neuromyelitis optica spectrum disorders (OR 1.292, 95% CI 1.043-1.543). ^58 60^ Additionally, COVID-19 had a positive effect on myasthenia gravis and celiac disease, with ORs of 1.096 and a 95% CI of 1.029-1.162^59^ and 1.111 and a 95% CI of 1.011-1.211^65^ (Figure 3). Conversely, COVID-19 had a negative effect with rheumatoid arthritis, with an OR of 0.976 and a 95% CI of 0.954-0.999.

In the context of the urogenital system, our meta-analysis indicated a positive association between COVID-19 and lower urinary tract symptoms, with an OR of 1.293 and a 95% CI of 1.043-1.543 (Figure 3).^71^

Furthermore, our study revealed a positive correlation between COVID-19 and various reproductive disorders.^71^ Specifically, there was a positive association between COVID-19 and sexual dysfunction, with an OR of 1.093 and a 95% CI of 1.026-1.16. This correlation suggests a heightened risk of sexual dysfunction in individuals with COVID-19 infection. COVID-19 was also positively linked to testicular dysfunction, with an OR of 1.326 and a 95% CI of 1.095-1.557. This finding implies a significantly increased risk of testicular dysfunction in the presence of COVID-19. In contrast, COVID-19 showed a negative correlation with torsion of the testis, with an OR of 0.772 and a 95% CI of 0.554-0.99 (Figure 3). This inverse relationship suggests that the risk of testicular torsion might be reduced in individuals with COVID-19.

## Discussion

This meta-analysis reveals a strong link between COVID-19 and heightened risks for multiple systemic diseases, including neuropsychiatric disorders (epilepsy, anxiety, schizophrenia), cerebrovascular diseases (stroke, thromboembolism), gastrointestinal issues (irritable bowel syndrome), endocrine diseases (type 2 diabetes), and autoimmune conditions (myasthenia gravis, neuromyelitis optica). The virus also impacts urinary and reproductive health, raising risks for lower urinary tract symptoms, erectile dysfunction, and testicular issues. Interestingly, COVID-19 infection seems to correlate with a lower likelihood of cognitive dysfunction, certain cardiac conditions (myocardial infarction, pericarditis), and testicular torsion.

It is well-established that angiotensin-converting enzyme 2 (ACE2) plays a pivotal role in both cardiovascular and immune system functions. Clinical studies have shown that SARS-CoV-2, the virus causing COVID-19, targets the ACE2 receptor to enter host cells, leading to acute myocardial injury and chronic cardiovascular complications.^97^ This can influence the progression or worsening of conditions such as stroke, diabetes, and myocardial infarction. ^98 99^ Additionally, a spectrum of neurological symptoms, including headaches, dizziness, altered consciousness, acute cerebrovascular events, ataxia, and epileptic seizures, have been documented post-COVID-19 infection. ^100^ Our research underscores that the consequences of COVID-19 extend beyond the acute phase, encompassing a wide array of long-term health impacts.

Our study has identified COVID-19 infection as a contributing factor in the onset or progression of epilepsy and stroke, aligning with previous clinical research that has observed alterations in mental state and cerebrovascular incidents among COVID-19 patients with neurological or psychiatric conditions.^101^ Despite some reports indicating that COVID-19 symptoms are predominantly respiratory or gastrointestinal and not typically associated with epileptic seizures,^102^ our research employs a Mendelian approach to infer causality using genetic variants as instrumental variables, thereby mitigating the impact of confounding factors. Our meta-research, in line with previous MR studies, consistently concludes that COVID-19 infection elevates the risk of epilepsy. Moreover, our findings reveal a statistically significant association between COVID-19 infection and cognitive disorders, optic nerve disorders, anxiety, and schizophrenia. However, given the limited number of studies included, further MR research is warranted to substantiate these findings. Regarding stroke, a meta-analysis has shown that patients with severe COVID-19 infection are at a higher risk of stroke.^103^ While this study discusses potential mechanisms for stroke and myocardial infarction in COVID-19 patients, the exact pathways remain to be elucidated.

### Strengths and Limitations

Our research boasts a number of significant strengths. It offers a comprehensive assessment of the causal links between COVID-19 and a spectrum of health outcomes, a contribution that is pioneering within the academic landscape. Adhering to the rigorous PRIMA and STROBE-MR guidelines, we’ve ensured a high standard of methodological integrity. The inclusion of a diverse array of studies from various databases and sources enhances both the robustness and the broad applicability of our findings. Most notably, our study delves into the genetic level impacts of COVID-19 on health, shedding light on its influence on disease susceptibility and severity, and offering fresh perspectives on the pandemic’s long-term health implications.

Our study, while insightful, has limitations. Aggregated data limits detailed patient insights, and a focus on European descendants may restrict the universality of our findings. Unmeasured confounders could affect associations, and we’ve assumed a linear relationship between COVID-19 and health outcomes, which may not capture all complexities. Future research should aim for large-scale, multicenter trials to solidify causal evidence between COVID-19 and health outcomes, providing a stronger basis for interventions and understanding long-term effects.

## Conclusion

In conclusion, our study systematically explored the causal relationships between COVID-19 and health outcomes across multiple human systems through meta-analysis of MR studies. We demonstrate that that COVID-19 increases the risk of diseases associated with the central nervous system and cardiovascular and cerebrovascular systems. Our study offers fresh insights, highlighting the need to delve into the long-term effects of COVID-19 and possible interventions. This research is crucial for a global evaluation of the pandemic’s impact and will lay the groundwork for strategies to alleviate the enduring health burden caused by COVID-19.

## Data Availability

All data produced in the present study are available upon reasonable request to the authors
All data produced in the present work are contained in the manuscript

## Contributors

LD, ZL, and YZ conceived and designed the study. LD, JW, MF, YY, XP, JJ, and HN acquired data. LD, JW, JJ, MD, and ZL analysed data. LD, JW, MF, YY, and XP interpreted the data. LD and YZ drafted the Article. ZL and MF critically revised the Article. All authors reviewed the submitted version of the manuscript and approved the final version of the manuscript. All authors had full access to all the data in the study and had final responsibility for the decision to submit for publication.

## Data sharing

All data included were derived from publicly available documents cited in the references. Extracted data are available upon request to the corresponding author.

## Acknowledgments

This work was funded by the Zhejiang Medical and Health Science and Technology Project (No.2024KY484), the Shaoxing Health Science and Technology Project (No.2023SKY073), the Science and Technology Planning Project of Shaoxing City (No. 2023A14030), and Shaoxing University Enterprise Important Horizontal Topic (No. 2024USXH287).

