## Supplemental 1 for "Causal associations of COVID-19 with health and disease outcomes: a systematic review and meta-analysis"

Supplementary material

eTable 1 Baseline characteristics of included studies in the meta-analysis.

| Study | Exposure  (A, B, C) | Study Source  of Exposure (HGI: Round) | Outcome | Cases（Outcome） | Controls（Outcome） | SNPs | OR (95% CI) - REM |
| --- | --- | --- | --- | --- | --- | --- | --- |
| Sun et al [2023] | C | R7 | AD | 71,880 | 383,378 | 20 | 1.0252 (0.9948 - 1.0565) |
| Sun et al [2023] | B | R7 | AD | 71,880 | 383,378 | 36 | 1.0163 (1.0045 - 1.0282) |
| Sun et al [2023] | A | R7 | AD | 71,880 | 383,378 | 31 | 1.0092 (1.0014 - 1.0171) |
| Ding et al [2023] | C | R7 | AD | 21,982 | 41,944 | 7 | 0.979 (0.808 - 1.03) |
| Ding et al [2023] | B | R7 | AD | 21,982 | 41,944 | 8 | 0.982 (0.9 - 1.07) |
| Ding et al [2023] | A | R7 | AD | 21,982 | 41,944 | 14 | 0.976 (0.924 - 1.03) |
| Ding et al [2023] | C | COVID-19-AncestryDNA GWAS | AD | 71,880 | 383,378 | 9 | 1.002 (0.998 - 1.005) |
| Ding et al [2023] | B | COVID-19-AncestryDNA GWAS | AD | 71,880 | 383,378 | 5 | 1 (0.997 - 1.004) |
| Ding et al [2023] | A | COVID-19-AncestryDNA GWAS | AD | 71,880 | 383,378 | 12 | 0.999 (0.998 - 1.001) |
| Zhang et al [2022] | C | R7 | AD | 954 | 487,331 | 4 | 1.0014 (1.0003 - 1.0025) |
| Zhang et al [2022] | B | R7 | AD | 954 | 487,331 | 4 | 1.0009 (1.00008 - 1.0017) |
| Zhang et al [2022] | A | R7 | AD | 954 | 487,331 | 4 | 1.0004 (1.00007 - 1.0008) |
| Zhang et al [2022] | C | R7 | ALS | 20,806 | 59,804 | 4 | 0.844 (0.802 - 0.888) |
| Zhang et al [2022] | B | R7 | ALS | 20,806 | 59,804 | 4 | 1.028 (0.913 - 1.157) |
| Zhang et al [2022] | A | R7 | ALS | 20,806 | 59,804 | 5 | 1.002 (0.937 - 1.724) |
| Zhang et al [2022] | C | R7 | MS | 47,429 | 68,374 | 4 | 0.786 (0.698 - 0.885) |
| Zhang et al [2022] | B | R7 | MS | 47,429 | 68,374 | 4 | 1.023 (0.907 - 1.155) |
| Zhang et al [2022] | A | R7 | MS | 47,429 | 68,374 | 5 | 1.001 (0.917 - 1.094) |
| Tirozzi et al [2022] | A | R6 | MDD | 170,756 | 329,443 | 17 | 0.999 (0.984 - 1.013) |
| Tirozzi et al [2022] | B | R6 | MDD | 170,756 | 329,443 | 18 | 0.995 (0.972 - 1.018) |
| Tirozzi et al [2022] | C | R6 | MDD | 170,756 | 329,443 | 17 | 0.972 (0.972 - 1.026) |
| Tirozzi et al [2022] | A | R6 | GAD | 16,730 | 101,021 | 20 | 1.006 (1 - 1.011) |
| Tirozzi et al [2022] | B | R6 | GAD | 16,730 | 101,021 | 23 | 1.01 (1.003 - 1.018) |
| Tirozzi et al [2022] | C | R6 | GAD | 16,730 | 101,021 | 19 | 0.956 (0.779 - 1.173) |
| Tirozzi et al [2022] | A | R6 | PD | 33,674 | 449,056 | 17 | 1.043 (0.885 - 1.23) |
| Tirozzi et al [2022] | B | R6 | PD | 33,674 | 449,056 | 19 | 1.104 (0.883 - 1.381) |
| Tirozzi et al [2022] | C | R6 | PD | 33,674 | 449,056 | 17 | 0.994 (0.977 - 1.026) |
| Tirozzi et al [2022] | A | R6 | SCZ | 35,476 | 46,839 | 19 | 1.021 (0.978 - 1.066) |
| Tirozzi et al [2022] | B | R6 | SCZ | 35,476 | 46,839 | 18 | 1.061 (0.996 - 1.129) |
| Tirozzi et al [2022] | C | R6 | SCZ | 35,476 | 46,839 | 17 | 0.768 (0.535 - 1.102) |
| Dai et al [2024] | C | R7 | AD | 8,835 | 205,799 | 23 | 1.01 (0.99 - 1.03) |
| Dai et al [2024] | B | R7 | AD | 8,835 | 205,799 | 28 | 1 (0.98 - 1.01)) |
| Dai et al [2024] | A | R7 | AD | 8,835 | 205,799 | 43 | 1 (0.99 - 1.02) |
| Dai et al [2024] | C | R7 | Benign meningioma | 9,660 | 445,127 | 22 | 0.96 (0.88 - 1.05) |
| Dai et al [2024] | B | R7 | Benign meningioma | 9,660 | 445,127 | 28 | 1 (0.93 - 1.07) |
| Dai et al [2024] | A | R7 | Benign meningioma | 9,660 | 445,127 | 41 | 1 (0.93 - 1.09) |
| Dai et al [2024] | C | R7 | Benign neoplasm of brain and other parts of CNS | 141 | 217,485 | 22 | 0.99 (0.89 - 1.09) |
| Dai et al [2024] | B | R7 | Benign neoplasm of brain and other parts of CNS | 141 | 217,485 | 28 | 1 (0.93 - 1.09)) |
| Dai et al [2024] | A | R7 | Benign neoplasm of brain and other parts of CNS | 141 | 217,485 | 41 | 1 (0.92 - 1.09) |
| Dai et al [2024] | C | R7 | Cerebral aneurysm | 26,888 | 7,037 | 24 | 1 (0.95 - 1.05) |
| Dai et al [2024] | B | R7 | Cerebral aneurysm | 26,888 | 7,037 | 31 | 0.99 (0.93 - 1.04) |
| Dai et al [2024] | A | R7 | Cerebral aneurysm | 26,888 | 7,037 | 44 | 0.98 (0.93 - 1.03) |
| Dai et al [2024] | C | R7 | Cerebral infarction | 7,264 | 49,373 | 23 | 1.02 (0.99 - 1.04) |
| Dai et al [2024] | B | R7 | Cerebral infarction | 7,264 | 49,373 | 30 | 1 (0.98 - 1.01) |
| Dai et al [2024] | A | R7 | Cerebral infarction | 7,264 | 49,373 | 43 | 1 (0.99 - 1.01) |
| Dai et al [2024] | C | R7 | Cervical spondylosis | 91 | 218,701 | 24 | 1 (0.99 - 1.01) |
| Dai et al [2024] | B | R7 | Cervical spondylosis | 91 | 218,701 | 29 | 1 (0.99 - 1.01)) |
| Dai et al [2024] | A | R7 | Cervical spondylosis | 91 | 218,701 | 43 | 1 (0.99 - 1.01) |
| Dai et al [2024] | C | R7 | Ceryical spinal cord and nerve injuries | 198 | 218,594 | 22 | 0.94 (0.78 - 1.14) |
| Dai et al [2024] | B | R7 | Ceryical spinal cord and nerve injuries | 198 | 218,594 | 28 | 0.98 (0.84 - 1.13) |
| Dai et al [2024] | A | R7 | Ceryical spinal cord and nerve injuries | 198 | 218,594 | 41 | 0.97 (0.82 - 1.13) |
| Dai et al [2024] | C | R7 | Concussion | 10,527 | 136,576 | 22 | 1.02 (0.98 - 1.05) |
| Dai et al [2024] | B | R7 | Concussion | 10,527 | 136,576 | 28 | 1.01 (0.99 - 1.03) |
| Dai et al [2024] | A | R7 | Concussion | 10,527 | 136,576 | 41 | 0.99 (0.96 - 1.02) |
| Dai et al [2024] | C | R7 | Congenital malformations of the nervous system | 258 | 218,534 | 22 | 1.14 (0.95 - 1.37) |
| Dai et al [2024] | B | R7 | Congenital malformations of the nervous system | 258 | 218,534 | 28 | 1.03 (0.87 - 1.22) |
| Dai et al [2024] | A | R7 | Congenital malformations of the nervous system | 258 | 218,534 | 41 | 1.11 (0.93 - 1.33) |
| Dai et al [2024] | C | R7 | Craniosynostosis | 405 | 218,387 | 22 | 0.99 (0.85 - 1.15) |
| Dai et al [2024] | B | R7 | Craniosynostosis | 405 | 218,387 | 28 | 1.02 (0.91 - 1.14) |
| Dai et al [2024] | A | R7 | Craniosynostosis | 405 | 218,387 | 41 | 0.95 (0.83 - 1.09) |
| Dai et al [2024] | C | R7 | Diffuse brain injury | 656 | 136,576 | 22 | 1.1 (0.95 - 1.29) |
| Dai et al [2024] | B | R7 | Diffuse brain injury | 656 | 136,576 | 28 | 1.06 (0.95 - 1.18) |
| Dai et al [2024] | A | R7 | Diffuse brain injury | 656 | 136,576 | 41 | 0.96 (0.87 - 1.07) |
| Dai et al [2024] | C | R7 | Epilepsy | 1,935 | 471,578 | 24 | 0.99 (0.95 - 1.03) |
| Dai et al [2024] | B | R7 | Epilepsy | 1,935 | 471,578 | 31 | 1 (0.97 - 1.04) |
| Dai et al [2024] | A | R7 | Epilepsy | 1,935 | 471,578 | 44 | 0.99 (0.95 - 1.03) |
| Dai et al [2024] | C | R7 | Focal brain injury | 1,065 | 136,576 | 22 | 1.05 (0.96 - 1.15) |
| Dai et al [2024] | B | R7 | Focal brain injury | 1,065 | 136,576 | 28 | 1.08 (1.00 - 1.18) |
| Dai et al [2024] | A | R7 | Focal brain injury | 1,065 | 136,576 | 41 | 0.99 (0.91 - 1.07) |
| Dai et al [2024] | C | R7 | Glioblastoma | 3,352 | 481,246 | 22 | 1.01 (0.74 - 1.38) |
| Dai et al [2024] | B | R7 | Glioblastoma | 3,352 | 481,246 | 28 | 1.08 (0.84 - 1.38) |
| Dai et al [2024] | A | R7 | Glioblastoma | 3,352 | 481,246 | 41 | 0.92 (0.69 - 1.23) |
| Dai et al [2024] | C | R7 | Hydrocephalus | 749 | 205,799 | 22 | 1.01 (0.90 - 1.12) |
| Dai et al [2024] | B | R7 | Hydrocephalus | 749 | 205,799 | 28 | 1 (0.91 - 1.10) |
| Dai et al [2024] | A | R7 | Hydrocephalus | 749 | 205,799 | 41 | 1.03 (0.93 - 1.13) |
| Dai et al [2024] | C | R7 | Intracerebral hemorrhage | 4,382 | 453,928 | 24 | 1 (0.95 - 1.05) |
| Dai et al [2024] | B | R7 | Intracerebral hemorrhage | 4,382 | 453,928 | 31 | 0.98 (0.93 - 1.03)) |
| Dai et al [2024] | A | R7 | Intracerebral hemorrhage | 4,382 | 453,928 | 44 | 0.98 (0.93 - 1.03) |
| Dai et al [2024] | C | R7 | Intracranial and intraspinal abscess | 923 | 217,869 | 22 | 1.07 (0.83 - 1.37) |
| Dai et al [2024] | B | R7 | Intracranial and intraspinal abscess | 923 | 217,869 | 28 | 1.06 (0.85 - 1.33) |
| Dai et al [2024] | A | R7 | Intracranial and intraspinal abscess | 923 | 217,869 | 41 | 1.14 (0.92 - 1.42) |
| Dai et al [2024] | C | R7 | MDD | 2,353 | 358,841 | 23 | 1.03 (0.97 - 1.08) |
| Dai et al [2024] | B | R7 | MDD | 2,353 | 358,841 | 24 | 1.03 (0.97 - 1.09) |
| Dai et al [2024] | A | R7 | MDD | 2,353 | 358,841 | 38 | 1.02 (0.96 - 1.08) |
| Dai et al [2024] | C | R7 | Malignant meningioma | 118 | 218,674 | 22 | 1.01 (0.90 - 1.13) |
| Dai et al [2024] | B | R7 | Malignant meningioma | 118 | 218,674 | 28 | 0.95 (0.86 - 1.05) |
| Dai et al [2024] | A | R7 | Malignant meningioma | 118 | 218,674 | 41 | 1.02 (0.92 - 1.13) |
| Dai et al [2024] | C | R7 | Malignant neoplasm of brain and other parts of CNS | 254 | 215,476 | 22 | 0.96 (0.77 - 1.18) |
| Dai et al [2024] | B | R7 | Malignant neoplasm of brain and other parts of CNS | 254 | 215,476 | 28 | 1.06 (0.88 - 1.27) |
| Dai et al [2024] | A | R7 | Malignant neoplasm of brain and other parts of CNS | 254 | 215,476 | 41 | 1.19 (0.97 - 1.47) |
| Dai et al [2024] | C | R7 | OCD | 945 | 472,738 | 23 | 1.02 (0.96 - 1.09) |
| Dai et al [2024] | B | R7 | OCD | 945 | 472,738 | 23 | 1.01 (0.93 - 1.10) |
| Dai et al [2024] | A | R7 | OCD | 945 | 472,738 | 38 | 1.02 (0.95 - 1.10) |
| Dai et al [2024] | C | R7 | PD | 1,693 | 471,562 | 24 | 0.98 (0.93 - 1.03) |
| Dai et al [2024] | B | R7 | PD | 1,693 | 471,562 | 31 | 1.03 (0.98 - 1.08) |
| Dai et al [2024] | A | R7 | PD | 1,693 | 471,562 | 44 | 0.98 (0.94 - 1.03) |
| Dai et al [2024] | C | R7 | Pituitary adenoma and craniopharyngioma | 183 | 164,682 | 22 | 1.04 (0.93 - 1.16) |
| Dai et al [2024] | B | R7 | Pituitary adenoma and craniopharyngioma | 183 | 164,682 | 28 | 0.9 (0.81 - 0.99) |
| Dai et al [2024] | A | R7 | Pituitary adenoma and craniopharyngioma | 183 | 164,682 | 41 | 1.03 (0.93 - 1.13) |
| Dai et al [2024] | C | R7 | Spinal canal stenosis | 1,147 | 217,645 | 24 | 1.01 (0.98 - 1.04) |
| Dai et al [2024] | B | R7 | Spinal canal stenosis | 1,147 | 217,645 | 31 | 0.98 (0.96 - 1.01) |
| Dai et al [2024] | A | R7 | Spinal canal stenosis | 1,147 | 217,645 | 44 | 1.03 (1.00 - 1.06) |
| Dai et al [2024] | C | R7 | Spinal meningioma | 640 | 218,152 | 22 | 0.85 (0.62 - 1.18) |
| Dai et al [2024] | B | R7 | Spinal meningioma | 640 | 218,152 | 28 | 0.83 (0.67 - 1.04) |
| Dai et al [2024] | A | R7 | Spinal meningioma | 640 | 218,152 | 41 | 1.21 (0.95 - 1.54) |
| Dai et al [2024] | C | R7 | Spinal osteochondrosis | 735 | 218,057 | 22 | 0.93 (0.75 - 1.16) |
| Dai et al [2024] | B | R7 | Spinal osteochondrosis | 735 | 218,057 | 28 | 1.02 (0.86 - 1.21) |
| Dai et al [2024] | A | R7 | Spinal osteochondrosis | 735 | 218,057 | 41 | 0.95 (0.78 - 1.17) |
| Dai et al [2024] | C | R7 | Stroke | 800 | 195,047 | 22 | 1 (0.99 - 1.03) |
| Dai et al [2024] | B | R7 | Stroke | 800 | 195,047 | 16 | 1.01 ( (0.99 - 1.03)) |
| Dai et al [2024] | A | R7 | Stroke | 800 | 195,047 | 39 | 1.02 (1.00 - 1.04) |
| Dai et al [2024] | C | R7 | Subarachnoid hemorrhage | 2,638 | 477,380 | 24 | 1.02 (0.96 - 1.07) |
| Dai et al [2024] | B | R7 | Subarachnoid hemorrhage | 2,638 | 477,380 | 31 | 0.98 (0.93 - 1.03)) |
| Dai et al [2024] | A | R7 | Subarachnoid hemorrhage | 2,638 | 477,380 | 44 | 1 (0.94 - 1.06) |
| Dai et al [2024] | C | R7 | TIA | 39,106 | 46,828 | 22 | 0.99 (0.95 - 1.02) |
| Dai et al [2024] | B | R7 | TIA | 39,106 | 46,828 | 28 | 1 (0.97 - 1.02)) |
| Dai et al [2024] | A | R7 | TIA | 39,106 | 46,828 | 41 | 1 (0.97 - 1.03) |
| Dai et al [2024] | C | R7 | Trigeminal ncuralgia | 40,585 | 406,111 | 22 | 1.05 (0.94 - 1.16) |
| Dai et al [2024] | B | R7 | Trigeminal ncuralgia | 40,585 | 406,111 | 28 | 1.02 (0.94 - 1.11) |
| Dai et al [2024] | A | R7 | Trigeminal ncuralgia | 40,585 | 406,111 | 41 | 1.05 (0.96 - 1.15) |
| Chen et al [2023] | C | R7 | ADHD | 38,691 | 275,986 | 26 | 0.99 (0.96 - 1.02) |
| Chen et al [2023] | B | R7 | ADHD | 38,691 | 275,986 | 26 | 1.00 (0.99 - 1.02) |
| Chen et al [2023] | A | R7 | ADHD | 38,691 | 275,986 | 26 | 1.00 (0.99 - 1.01) |
| Chen et al [2023] | C | R7 | ASD | 18,381 | 27,969 | 36 | 0.94 (0.81 - 1.08) |
| Chen et al [2023] | B | R7 | ASD | 18,381 | 27,969 | 36 | 0.99 (0.93 - 1.05) |
| Chen et al [2023] | A | R7 | ASD | 18,381 | 27,969 | 36 | 0.99 (0.96 - 1.03) |
| Chen et al [2023] | C | R7 | TS | 4,819 | 9,488 | 58 | 1.01 (0.94 - 1.09) |
| Chen et al [2023] | B | R7 | TS | 4,819 | 9,488 | 57 | 0.99 (0.95 - 1.02) |
| Chen et al [2023] | A | R7 | TS | 4,819 | 9,488 | 56 | 1.00 (0 97 - 1.02) |
| Baranova et al [2022] | C | R4/R5 | SCZ | 53,386 | 77,258 | 20 | 1.06 (0.83 - 1.37) |
| Baranova et al [2022] | B | R4/R5 | SCZ | 53,386 | 77,258 | 36 | 1.11 (1.02 - 1.20) |
| Ran et al [2023] | C | R7 | Cognitive dysfunction | NA | NA | 20 | 0.921 (0.876 - 0.969) |
| Ran et al [2023] | B | R7 | Cognitive dysfunction | NA | NA | 36 | 0.832 (0.743 - 0.933) |
| Ran et al [2023] | A | R7 | Cognitive dysfunction | NA | NA | 32 | 0.780 (0.655 - 0.929) |
| Cao et al [2023] | C | R7 | Intelligence | 32,519 | 2,062,805 | 19 | 0.965 (0.939 - 0.993) |
| Cao et al [2023] | B | R7 | Intelligence | 32,519 | 2,062,805 | 41 | 0.988 (0.972 - 1.003) |
| Cao et al [2023] | A | R7 | Intelligence | 32,519 | 2,062,805 | 34 | 0.988 (0.972 - 1.003) |
| Zhu et al [2022] | B | R6 | Childhood Intelligence | NA | NA | 4 | 0.859 (0.776 - 0.952) |
| Ran et al [2023] | C | R7 | Insomnia | NA | NA | 16 | 1.011 (0.988 - 1.035) |
| Ran et al [2023] | B | R7 | Insomnia | NA | NA | 39 | 0.997 (0.989 - 1.005) |
| Ran et al [2023] | A | R7 | Insomnia | NA | NA | 34 | 0.996 (0.991 - 1.001) |
| Li et al [2023] | C | R7 | MDD | 65,075 | 232,552 | 8 | 0.974 (0.938 - 1.011) |
| Li et al [2023] | B | R7 | MDD | 65,075 | 232,552 | 9 | 0.985 (0.966 - 1.005) |
| Li et al [2023] | A | R7 | MDD | 65,075 | 232,552 | 4 | 0.999 (0.984 - 1.014) |
| Baranova et al [2023] | C | R7 | MDD | 246,363 | 561,190 | 20 | 1.01 (0.97 - 1.05) |
| Baranova et al [2023] | B | R7 | MDD | 246,363 | 561,190 | 36 | 1 (0.98 - 1.02) |
| Baranova et al [2023] | A | R7 | MDD | 246,363 | 561,190 | 38 | 1 (0.99 - 1.01) |
| Xue et al [2023] | C | R7 | ASD | 18,382 | 27,969 | 3 | 0.971 (0.926 - 1.019) |
| Xue et al [2023] | B | R7 | ASD | 18,382 | 27,969 | 6 | 0.982 (0.903 - 1.068) |
| Xue et al [2023] | A | R7 | ASD | 18,382 | 27,969 | 6 | 0.994 (0.933 - 1.059) |
| Xue et al [2023] | C | R7 | MDD | 135,458 | 344,901 | 5 | 1.044 (0.949 - 1.149) |
| Xue et al [2023] | B | R7 | MDD | 135,458 | 344,901 | 6 | 1.036 (0.990 - 1.084) |
| Xue et al [2023] | A | R7 | MDD | 135,458 | 344,901 | 6 | 1.002 (0.996 - 1.008) |
| Xue et al [2023] | C | R7 | BD | 20,352 | 31,358 | 5 | 0.894 (0.707 - 1.130) |
| Xue et al [2023] | B | R7 | BD | 20,352 | 31,358 | 7 | 1.32 (1.106 - 1.576) |
| Xue et al [2023] | A | R7 | BD | 20,352 | 31,358 | 7 | 1.139 (1.033 - 1.256) |
| Xue et al [2023] | C | R7 | SCZ | 33,640 | 43,456 | 5 | 0.971 (0.847 - 1.114) |
| Xue et al [2023] | B | R7 | SCZ | 33,640 | 43,456 | 6 | 1.096 (1.031 - 1.164) |
| Xue et al [2023] | A | R7 | SCZ | 33,640 | 43,456 | 7 | 1.043 (1.005 - 1.082) |
| Xue et al [2023] | C | R7 | GAD | 25,453 | 58,113 | 3 | 1.001 (0.999 - 1.002) |
| Xue et al [2023] | B | R7 | GAD | 25,453 | 58,113 | 6 | 1.043 (0.977 - 1.114) |
| Xue et al [2023] | A | R7 | GAD | 25,453 | 58,113 | 6 | 1.01 (0.961 - 1.061) |
| Pan et al [2024] | C | R7 | mRNFL | NA | NA | 22 | 1.74 (1.20 - 2.52) |
| Pan et al [2024] | B | R7 | mRNFL | NA | NA | 40 | 1.11 (0.90 - 1.37) |
| Pan et al [2024] | C | R7 | mGCIPL | NA | NA | 22 | 2.43 (1.49 - 3.96) |
| Pan et al [2024] | B | R7 | mGCIPL | NA | NA | 40 | 1.28 (0.88 - 1.85) |
| Xing et al [2023] | C | R7 | Pre-eclampsia | 3,903 | 114,735 | 5 | 1.23 (1.01 - 1.51) |
| Xing et al [2023] | B | R7 | Pre-eclampsia | 3,903 | 114,735 | 5 | 1.15 (1.02 - 1.3) |
| Xing et al [2023] | C | R7 | Eclampsia | 3,556 | 114,735 | 6 | 1.22 (0.99 - 1.51) |
| Xing et al [2023] | B | R7 | Eclampsia | 3,556 | 114,735 | 5 | 1.14 (1.01 - 1.3) |
| You et al [2023] | C | FinnGen | Epilepsy | 10,354 | 264,662 | 8 | 1.5306 (1.1676 - 2.0062) |
| You et al [2023] | B | FinnGen | Epilepsy | 10,354 | 264,662 | 15 | 1.0934 (1.0097 - 1.1841) |
| You et al [2023] | A | FinnGen | Epilepsy | 10,354 | 264,662 | 9 | 1.2454 (1.085 - 1.4295) |
| You et al [2023] | C | FinnGen | Epilepsy | 15,212 | 29,677 | 3 | 1.344 (1.0235 - 1.7649) |
| You et al [2023] | B | FinnGen | Epilepsy | 15,212 | 29,677 | 3 | 1.7381 (1.0467 - 2.8862) |
| You et al [2023] | A | FinnGen | Epilepsy | 15,212 | 29,677 | 4 | 1.2724 (1.0347 - 1.5647) |
| You et al [2023] | C | FinnGen | Focal epilepsy | 1,132 | 332,145 | 6 | 1.0161 (0.6269 - 1.498) |
| You et al [2023] | B | FinnGen | Focal epilepsy | 1,132 | 332,145 | 9 | 1.5846 (1.0099 - 2.4863) |
| You et al [2023] | A | FinnGen | Focal epilepsy | 1,132 | 332,145 | 20 | 1.6818 (1.1478 - 2.4642) |
| You et al [2023] | C | FinnGen | Focal epilepsy | 9,671 | 29,677 | 7 | 1.0869 (0.9715 - 1.2159) |
| You et al [2023] | B | FinnGen | Focal epilepsy | 9,671 | 29,677 | 8 | 1.7549 (1.1063 - 2.7838) |
| You et al [2023] | A | FinnGen | Focal epilepsy | 9,671 | 29,677 | 8 | 1.6598 (1.2572 - 2.1914) |
| You et al [2023] | C | FinnGen | Generalized epilepsy | 1,988 | 332,145 | 9 | 2.1155 (1.1734 - 3.8139) |
| You et al [2023] | B | FinnGen | Generalized epilepsy | 1,988 | 332,145 | 7 | 1.2281 (1.0513 - 1.4346) |
| You et al [2023] | A | FinnGen | Generalized epilepsy | 1,988 | 332,145 | 20 | 1.1486 (1.0274 - 1.2842) |
| You et al [2023] | C | FinnGen | Generalized epilepsy | 3,769 | 29,677 | 8 | 1.1245 (1.0444 - 1.2108) |
| You et al [2023] | B | FinnGen | Generalized epilepsy | 3,769 | 29,677 | 6 | 1.1827 (1.0215 - 1.3693) |
| You et al [2023] | A | FinnGen | Generalized epilepsy | 3,769 | 29,677 | 6 | 1.0439 (1.0159 - 1.0728) |
| He et al [2023] | C | R7 | Epilepsy | 15,212 | 29,677 | 7 | 0.99 (0.86 - 1.14) |
| He et al [2023] | B | R7 | Epilepsy | 15,212 | 29,677 | 19 | 1 (0.96 - 1.04) |
| He et al [2023] | A | R7 | Epilepsy | 15,212 | 29,677 | 19 | 0.99 (0.96 - 1.01) |
| Wu et al [2024] | A | R5 | pre-eclampsia | 3,556 | 114,735 | 7 | 1.10 (1.01 - 1.19) |
| Wu et al [2024] | B | R5 | pre-eclampsia | 3,556 | 114,735 | 6 | 1.17 (1.03 - 1.33) |
| Wu et al [2024] | C | R5 | pre-eclampsia | 3,556 | 114,735 | 7 | 1.15 (0.88 - 1.52) |
| Wu et al [2024] | A | R5 | eclampsia | 290 | 114,735 | 7 | 0.93 (0.69 - 1.25) |
| Wu et al [2024] | B | R5 | eclampsia | 290 | 114,735 | 6 | 0.90 (0.61 - 1.31) |
| Wu et al [2024] | C | R5 | eclampsia | 290 | 114,735 | 7 | 0.44 (0.18 - 1.11) |
| Baranova et al [2024] | B | R7 | PTSD | 23,212 | 151,447 | 33 | 1.00 (0.98 - 1.01) |
| Baranova et al [2024] | A | R7 | PTSD | 23,212 | 151,447 | 14 | 0.99 (0.95 - 1.03) |
| Cao et al [2024] | C | R5 | Optic nerve and visual pathway disorders | 1,301 | 217,491 | 9 | 1.697 (1.086 - 2.652) |
| Jiang et al [2024] | A | R7 | Migraine | 1,072 | 360,122 | 40 | 1.0000881 (0.999748 - 1.000428) |
| Jiang et al [2024] | B | R7 | Migraine | 1,072 | 360,122 | 43 | 1.000024 (0.9994893 - 1.000559) |
| Jiang et al [2024] | C | R7 | Migraine | 1,072 | 360,122 | 23 | 1.000358 (0.999023 - 1.001695) |
| Monistrol-Mula et al [2024] | B | R7 | MDD | 371,184 | 978,703 | 38 | 1.00 (0.98 - 1.01) |
| Monistrol-Mula et al [2024] | C | R7 | MDD | 371,184 | 978,703 | 23 | 0.99 (0.95 - 1.05) |
| Monistrol-Mula et al [2024] | B | R7 | BD | 41,917 | 371,549 | 38 | 1.02 (0.97 - 1.07) |
| Monistrol-Mula et al [2024] | C | R7 | BD | 41,917 | 371,549 | 23 | 1.05 (0.90 - 1.22) |
| Monistrol-Mula et al [2024] | B | R7 | PTSD | 137,136 | 1,085,746 | 38 | 1.00 (0.99 - 1.00) |
| Monistrol-Mula et al [2024] | C | R7 | PTSD | 137,136 | 1,085,746 | 23 | 0.99 (0.97 - 1.02) |
| Tan et al [2021] | A | GenOMICC | Pregnancy-induced hypertension | NA | NA | 8 | 1.111 (1.042 - 1.184) |
| Zhang et al [2022] | C | R5 | AF | 18,398 | 91,536 | 29 | 1.111 (0.971 - 1.272) |
| Zhang et al [2022] | B | R5 | AF | 18,398 | 91,536 | 20 | 0.991 (0.941 - 1.044) |
| Zhang et al [2022] | B | R5 | AF | 18,398 | 91,536 | 32 | 1.055 (0.995 - 1.119) |
| Zhang et al [2022] | A | R5 | AF | 18,398 | 91,536 | 33 | 1.037 (1.005 - 1.071) |
| Tan et al [2022] | B | R5 | AF | 55,114 | 482,295 | NA | 1.004 (0.935 - 1.061) |
| Tan et al [2022] | A | R5 | AF | 55,115 | 482,296 | NA | 0.992 (0.954 - 1.032) |
| Tan et al [2022] | B | R5(without UKBB data) | AF | 55,116 | 482,297 | NA | 0.999 (0.624 - 1.599) |
| Tan et al [2022] | A | R5(without UKBB data) | AF | 55,117 | 482,298 | NA | 0.999 (0.962 - 1.037) |
| Tan et al [2022] | B | R5 | CAD | 34,541 | 261,984 | NA | 1.007 (0.955 - 1.062) |
| Tan et al [2022] | A | R5 | CAD | 34,541 | 261,984 | NA | 1.004 (0.975 - 1.034) |
| Tan et al [2022] | B | R5(without UKBB data) | CAD | 34,541 | 261,984 | NA | 1.006 (0.962 - 1.052) |
| Tan et al [2022] | A | R5(without UKBB data) | CAD | 34,541 | 261,984 | NA | 1.007 (0.974 - 1.041) |
| Zuber et al [2021] | C | R5 | CES | 4,373 | 406,111 | 31 | 1.06 (1.01 - 1.12) |
| Zuber et al [2021] | C | R5 | LAS | 7,193 | 406,111 | 31 | 1.07 (0.997 - 1.141) |
| Zuber et al [2021] | C | R5 | SVS | 5,386 | 406,111 | 31 | 1.054 (0.999 - 1.112) |
| Zhang et al [2022] | C | R6 | Stroke | 40,585 | 406,111 | 8 | 1.03 (0.96 - 1.11) |
| Zhang et al [2022] | B | R6 | Stroke | 40,585 | 406,111 | 18 | 1.06 (0.96 - 1.21) |
| Zhang et al [2022] | A | R6 | Stroke | 40,585 | 406,111 | 15 | 1.05 (1.01 - 1.1) |
| Zhang et al [2022] | C | R6 | IS | 34,217 | 406,111 | 8 | 1.06 (1.02 - 1.11) |
| Zhang et al [2022] | B | R6 | IS | 34,217 | 406,111 | 18 | 1.04 (1.01 - 1.06) |
| Zhang et al [2022] | A | R6 | IS | 34,217 | 406,111 | 15 | 1.06 (1.02 - 1.09) |
| Huang et al [2023] | A | R7 | VTE | 3,900 | 369,592 | 98 | 1.11 (1.06 - 1.17) |
| Huang et al [2023] | B | R7 | VTE | 3,900 | 369,592 | 99 | 1.10 (1.06 - 1.14) |
| Huang et al [2023] | C | R7 | VTE | 3,900 | 369,592 | 102 | 1.06 (1.04 - 1.09) |
| Wang et al [2022] | C | R5 | HF | 47,309 | 930,014 | 5 | 1.066 (0.955 - 1.19) |
| Wang et al [2022] | B | R5 | HF | 47,309 | 930,014 | 5 | 1.009 (0.939 - 1.085) |
| Wang et al [2022] | A | R5 | HF | 47,309 | 930,014 | 9 | 1.003 (0.969 - 1.037) |
| Zheng et al [2023] | A | R7 | MI | NA | NA | 26 | 0.992 (0.97 - 1.014) |
| Zheng et al [2023] | C | R7 | MI | NA | NA | 14 | 1.004 (0.923 - 1.092) |
| Zheng et al [2023] | A | R7 | MI | NA | NA | 28 | 0.964 (0.936 - 0.993) |
| Zheng et al [2023] | C | R7 | MI | NA | NA | 14 | 0.965 (0.88 - 1.06) |
| Zheng et al [2023] | A | R7 | MI | NA | NA | 28 | 0.973 (0.936 - 1.012) |
| Zheng et al [2023] | C | R7 | MI | NA | NA | 14 | 1.07 (0.935 - 1.224) |
| Wang et al [2023] | A | R5 | CHD | 22,233 | 64,762 | NA | 1.01 (1.01 - 1.02) |
| Jia et al [2022] | A | GenOMICC | Aortic Aneurysms | 791 | 89,376 | 8 | 1.066 (0.939 - 1.211) |
| Jia et al [2022] | A | GenOMICC | AF | 7,244 | 56,378 | 8 | 0.982 (0.928 - 1.040) |
| Jia et al [2022] | A | GenOMICC | VTE | 1,785 | 84,462 | 8 | 1.026 (0.921 - 1.143) |
| Jia et al [2022] | A | GenOMICC | HF | 8,016 | 75,137 | 8 | 1.049 (1.001 - 1.100) |
| Jia et al [2022] | A | GenOMICC | HTN | 22,154 | 56,378 | 8 | 1.020 (0.977 - 1.065) |
| Jia et al [2022] | A | GenOMICC | Major coronary heart disease event | 7,123 | 89,376 | 8 | 1.081 (1.007 - 1.160) |
| Jia et al [2022] | A | GenOMICC | NRVHD | 3,108 | 75,137 | 8 | 1.010 (0.936 - 1.090) |
| Jia et al [2022] | A | GenOMICC | PAD | 2,383 | 92,349 | 8 | 1.059 (0.905 - 1.238) |
| Jia et al [2022] | A | GenOMICC | PE | 1,366 | 95,023 | 8 | 0.989 (0.898 - 1.089) |
| Jia et al [2022] | A | GenOMICC | RHD | 162 | 96,273 | 8 | 1.023 (0.778 - 1.346) |
| Liu et al [2023] | A | R7 | Myocarditis | 1,521 | 191,924 | 21 | 1.00 (0.89 - 1.12) |
| Liu et al [2023] | B | R7 | Myocarditis | 1,521 | 191,924 | 27 | 0.98 (0.84 - 1.15) |
| Liu et al [2023] | C | R7 | Myocarditis | 1,521 | 191,924 | 10 | 0.98 (0.84 - 1.15) |
| Liu et al [2023] | A | R7 | Pericarditis | 979 | 286,109 | 21 | 0.90 (0.78 - 1.04) |
| Liu et al [2023] | B | R7 | Pericarditis | 979 | 286,109 | 27 | 0.90 (0.78 - 1.04) |
| Liu et al [2023] | C | R7 | Pericarditis | 979 | 286,109 | 11 | 0.90 (0.78 - 1.04) |
| Ma et al [2023] | A | R7 | IS | 62,100 | 1,234,808 | 27 | 1.01 (0.98 - 1.05) |
| Ma et al [2023] | A | R7 | CES | 10,804 | 234,808 | 26 | 1.07 (1.01 - 1.14) |
| Ma et al [2023] | A | R7 | LAS | 6,399 | 1,234,808 | 25 | 1.04 (0.93 - 1.16) |
| Ma et al [2023] | A | R7 | SVS | 68,111 | 234,808 | 23 | 1.00 (0.94 - 1.06) |
| Li et al [2024] | C | R5 | VTE | 8,077 | 303,091 | 6 | 1.02 (0.83 - 1.26) |
| Li et al [2024] | B | R5 | VTE | 8,077 | 303,091 | 4 | 1.03 (0.94 - 1.13) |
| Li et al [2024] | A | R5 | VTE | 8,077 | 303,091 | 7 | 0.99 (0.92 - 1.07) |
| Wang et al [2024] | A | R7 | VTE | 19,372 | 201,194 | 26 | 0.974 (0.936 - 1.013) |
| Wang et al [2024] | B | R7 | VTE | 19,372 | 201,194 | 31 | 0.976 (0.918 - 1.039) |
| Wang et al [2024] | C | R7 | VTE | 19,372 | 201,194 | 13 | 0.908 (0.775 - 1.065) |
| Tang et al [2024] | A | R7 | Cardiac arrest | 3,939 | 25,989 | 9 | 0.99 (0.81 - 1.21) |
| Tang et al [2024] | B | R7 | Cardiac arrest | 3,939 | 25,989 | 6 | 1.02 (0.70 - 1.49) |
| Tang et al [2024] | C | R7 | Cardiac arrest | 3,939 | 25,989 | 6 | 1.12 (0.47 - 2.67) |
| Tang et al [2024] | A | R7 | Cardiac arrest | 2,471 | 210,652 | 22 | 0.94 (0.85 - 1.03) |
| Tang et al [2024] | B | R7 | Cardiac arrest | 2,471 | 210,652 | 21 | 0.98 (0.85 - 1.12) |
| Tang et al [2024] | C | R7 | Cardiac arrest | 2,471 | 210,652 | 11 | 1.20 (0.75 - 1.90) |
| Li et al [2024] | C | R5 | Myocarditis | 829 | 217,963 | 7 | 1.407 (0.761 - 2.602) |
| Li et al [2024] | C | R5 | AMI | 3,927 | 333,272 | 7 | 1.002 (0.999 - 1.005) |
| Li et al [2024] | C | R5 | Arrhythmia | 17,861 | 194,592 | 6 | 0.865 (0.717 - 1.044) |
| Li et al [2024] | C | R5 | VTE | 4,620 | 356,574 | 7 | 1.013 (0.997 - 1.028) |
| Ong et al [2021] | A | R5 | GERD | 78,707 | 288,734 | 6 | 1.014 (0.996 - 1.033) |
| Ong et al [2021] | B | R5 | GERD | 78,707 | 288,734 | 5 | 1.016 (0.994 - 1.04) |
| Ong et al [2021] | C | R5 | GERD | 78,707 | 288,734 | 3 | 0.953 (0.902 - 1.008) |
| Wang et al [2023] | C | R5 | Acute Appendicitis | 11,899 | 201,886 | 7 | 0.93 (0.85 - 1.02) |
| Wang et al [2023] | B | R5 | Acute Appendicitis | 11,899 | 201,886 | 5 | 1.36 (0.82 - 2.25) |
| Wang et al [2023] | A | R5 | Acute Appendicitis | 11,899 | 201,886 | 8 | 1.05 (0.82 - 1.35) |
| Wang et al [2023] | C | R5 | AG | 1,284 | 189,695 | 7 | 1 (1 - 1) |
| Wang et al [2023] | B | R5 | AG | 1,284 | 189,695 | 5 | 1.0 (0.99 - 1) |
| Wang et al [2023] | A | R5 | AG | 1,284 | 189,695 | 8 | 1 (0.1 - 1) |
| Wang et al [2023] | C | R5 | AP | 3022 | 195,144 | 7 | 1.02 (0.98 - 1.06) |
| Wang et al [2023] | B | R5 | AP | 3022 | 195,144 | 5 | 1.16 (0.81 - 1.66) |
| Wang et al [2023] | A | R5 | AP | 3022 | 195,144 | 8 | 0.96 (0.77 - 1.18) |
| Wang et al [2023] | C | R5 | Chlocystitis | 2,013 | 195,144 | 7 | 0.94 (0.79 - 1.12) |
| Wang et al [2023] | B | R5 | Chlocystitis | 2,013 | 195,144 | 5 | 1.30 (0.98 - 1.72) |
| Wang et al [2023] | A | R5 | Chlocystitis | 2,013 | 195,144 | 8 | 1.07 (0.91 - 1.26) |
| Wang et al [2023] | C | R5 | Cholangitis | 778 | 195,144 | 7 | 0.92 (0.85 - 1) |
| Wang et al [2023] | B | R5 | Cholangitis | 778 | 195,144 | 5 | 0.96 (0.54 - 1.73) |
| Wang et al [2023] | A | R5 | Cholangitis | 778 | 195,144 | 8 | 1.18 (0.91 - 1.53) |
| Wang et al [2023] | C | R5 | Cholelithiasis | 19,023 | 195,144 | 7 | 1.02 (0.85 - 1.22) |
| Wang et al [2023] | B | R5 | Cholelithiasis | 19,023 | 195,144 | 5 | 1.36 (0.78 - 2.37) |
| Wang et al [2023] | A | R5 | Cholelithiasis | 19,023 | 195,144 | 8 | 0.91 (0.7 - 1.19) |
| Wang et al [2023] | C | R5 | Chronic Gastritis | 5213 | 189,695 | 7 | 1 (0.1 - 1) |
| Wang et al [2023] | B | R5 | Chronic Gastritis | 5213 | 189,695 | 5 | 0.87 (0.54 - 1.4) |
| Wang et al [2023] | A | R5 | Chronic Gastritis | 5213 | 189,695 | 8 | 0.97 (0.79 - 1.2) |
| Wang et al [2023] | C | R5 | CP | 1,737 | 195,144 | 7 | 0.93 (0.83 - 1.03) |
| Wang et al [2023] | B | R5 | CP | 1,737 | 195,144 | 5 | 0.99 (0.62 - 1.58) |
| Wang et al [2023] | A | R5 | CP | 1,737 | 195,144 | 8 | 0.97 (0.76 - 1.24) |
| Wang et al [2023] | C | R5 | CRC | 5,657 | 372,016 | 7 | 1.01 (0.97 - 1.06) |
| Wang et al [2023] | B | R5 | CRC | 5,657 | 372,016 | 5 | 1.00 (1 - 1.1) |
| Wang et al [2023] | A | R5 | CRC | 5,657 | 372,016 | 8 | 1 (1 - 1.1) |
| Wang et al [2023] | C | R5 | Crohn | 732 | 336,467 | 7 | 1.00 (0.89 - 1.12) |
| Wang et al [2023] | B | R5 | Crohn | 732 | 336,467 | 5 | 1.09 (0.94 - 1.27) |
| Wang et al [2023] | A | R5 | Crohn | 732 | 336,467 | 8 | 1.05 (0.97 - 1.12) |
| Wang et al [2023] | C | R5 | DL | 1,908 | 461,025 | 4 | 1 (1 - 1) |
| Wang et al [2023] | B | R5 | DL | 1,908 | 461,025 | 4 | 1 (1 - 1) |
| Wang et al [2023] | A | R5 | DL | 1,908 | 461,025 | 3 | 1 (1 - 1) |
| Wang et al [2023] | C | R5 | Gastric carcinoma | 6,563 | 195,745 | 6 | 1.00 (1 - 1.1) |
| Wang et al [2023] | B | R5 | Gastric carcinoma | 6,563 | 195,745 | 4 | 1 (1 - 1) |
| Wang et al [2023] | A | R5 | Gastric carcinoma | 6,563 | 195,745 | 7 | 0.94 (0.86 - 1.03) |
| Wang et al [2023] | C | R5 | GERD | 129,080 | 473,524 | 4 | 1.09 (1.01 - 1.18) |
| Wang et al [2023] | B | R5 | GERD | 129,080 | 473,524 | 4 | 1 (1 - 1) |
| Wang et al [2023] | A | R5 | GERD | 129,080 | 473,524 | 5 | 1 (1 - 1) |
| Wang et al [2023] | C | R5 | GS | 1,834 | 35,936 | 7 | 1 (1 - 1) |
| Wang et al [2023] | B | R5 | GS | 1,834 | 35,936 | 5 | 1 (1 - 1) |
| Wang et al [2023] | A | R5 | GS | 1,834 | 35,936 | 7 | 1.02 (1 - 1.04) |
| Wang et al [2023] | C | R5 | IBS | 10,939 | 451,994 | 7 | 0.95 (0.55 - 1.64) |
| Wang et al [2023] | B | R5 | IBS | 10,939 | 451,994 | 5 | 1 (0.86 - 1.16) |
| Wang et al [2023] | A | R5 | IBS | 10,939 | 451,994 | 7 | 1.04 (1.01 - 1.06) |
| Wang et al [2023] | C | R5 | NAFLD | 894 | 217,898 | 7 | 0.98 (0.89 - 1.08) |
| Wang et al [2023] | B | R5 | NAFLD | 894 | 217,898 | 5 | 1.11 (0.85 - 1.5) |
| Wang et al [2023] | A | R5 | NAFLD | 894 | 217,898 | 8 | 0.96 (0.79 - 1.16) |
| Wang et al [2023] | C | R5 | UC | 4,320 | 210,300 | 7 | 0.95 (0.83 - 1.09) |
| Wang et al [2023] | B | R5 | UC | 4,320 | 210,300 | 5 | 1.24 (0.99 - 1.6) |
| Wang et al [2023] | A | R5 | UC | 4,320 | 210,300 | 8 | 1.06 (0.92 - 1.22) |
| Ge et al [2023] | C | R5 | AAP | 931 | 376,346 | 31 | 0.968 (0.89 - 1.06) |
| Ge et al [2023] | B | R5 | AAP | 931 | 376,346 | 32 | 0.887 (0.78 - 1.01) |
| Ge et al [2023] | B | R5 | AAP | 931 | 376,346 | 25 | 1.013 (0.96 - 1.07) |
| Ge et al [2023] | A | R5 | AAP | 931 | 376,346 | 15 | 0.925 (0.81 - 1.05) |
| Ge et al [2023] | A | R5 | AAP | 931 | 376,346 | 42 | 1.036 (0.97 - 1.11) |
| Ge et al [2023] | A | R5 | AAP | 931 | 376,346 | 23 | 1.077 (0.98 - 1.18) |
| Ge et al [2023] | C | R5 | ACP | 1,794 | 375,483 | 31 | 0.985 (0.93 - 1.05) |
| Ge et al [2023] | B | R5 | ACP | 1,794 | 375,483 | 32 | 0.94 (0.87 - 1.02) |
| Ge et al [2023] | B | R5 | ACP | 1,794 | 375,483 | 25 | 0.993 (0.95 - 1.03) |
| Ge et al [2023] | A | R5 | ACP | 1,794 | 375,483 | 15 | 1.012 (0.91 - 1.12) |
| Ge et al [2023] | A | R5 | ACP | 1,794 | 375,483 | 42 | 0.992 (0.93 - 1.05) |
| Ge et al [2023] | A | R5 | ACP | 1,794 | 375,483 | 22 | 0.914 (0.85 - 0.98) |
| Ge et al [2023] | C | R5 | AP | 6,223 | 330,903 | 31 | 1.01 (0.88 - 1.16) |
| Ge et al [2023] | B | R5 | AP | 6,223 | 330,903 | 32 | 0.724 (0.51 - 1.03) |
| Ge et al [2023] | B | R5 | AP | 6,223 | 330,903 | 25 | 0.891 (0.74 - 1.07) |
| Ge et al [2023] | A | R5 | AP | 6,223 | 330,903 | 15 | 0.905 (0.62 - 1.21) |
| Ge et al [2023] | A | R5 | AP | 6,223 | 330,903 | 42 | 1.03 (0.91 - 1.11) |
| Ge et al [2023] | A | R5 | AP | 6,223 | 330,903 | 23 | 0.952 (0.81 - 1.12) |
| Ge et al [2023] | C | R5 | CP | 3,320 | 330,903 | 31 | 1.008 (0.97 - 1.04) |
| Ge et al [2023] | B | R5 | CP | 3,320 | 330,903 | 32 | 1.024 (0.93 - 1.13) |
| Ge et al [2023] | B | R5 | CP | 3,320 | 330,903 | 25 | 1.015 (0.97 - 1.07) |
| Ge et al [2023] | A | R5 | CP | 3,320 | 330,903 | 15 | 0.987 (0.92 - 1.06) |
| Ge et al [2023] | A | R5 | CP | 3,320 | 330,903 | 42 | 0.98 (0.94 - 1.02) |
| Ge et al [2023] | A | R5 | CP | 3,320 | 330,903 | 22 | 1.001 (0.91 - 1.1) |
| Liu et al [2021] | C | R6 | NAFLD | 1,687 | 398,277 | 7 | 1.054 (0.696 - 1.596) |
| Liu et al [2021] | B | R6 | NAFLD | 1,687 | 398,277 | 10 | 0.973 (0.750 - 1.263) |
| Liu et al [2021] | A | R5 | NAFLD | 1,687 | 398,277 | 8 | 0.919 (0.839 - 1.007) |
| He et al [2024] | A | R7 | Barrett's esophagus | 946 | 292256 | 32 | 1.06 (0.92 - 1.21) |
| He et al [2024] | A | R7 | Perforation of esophagus | 103 | 292256 | 32 | 1.07 (0.71 - 1.62) |
| He et al [2024] | A | R7 | Oesophageal obstruction | 879 | 292256 | 33 | 0.92 (0.81 - 1.06) |
| He et al [2024] | A | R7 | Esophageal varices | 894 | 295014 | 32 | 1.09 (0.95 - 1.24) |
| He et al [2024] | A | R7 | Esophageal ulcer | 1840 | 292256 | 32 | 0.99 (0.89 - 1.10) |
| He et al [2024] | A | R7 | Benign esophageal tumors | 260 | 342239 | 32 | 0.88 (0.69 - 1.13) |
| He et al [2024] | A | R7 | Esophagitis | 1318 | 292256 | 32 | 1.06 (0.94 - 1.19) |
| He et al [2024] | A | R7 | Gastroesophageal reflux disease | 22867 | 292256 | 33 | 0.99 (0.96 - 1.02) |
| He et al [2024] | A | R7 | Congenital esophageal malformation | 94 | 341501 | 32 | 1.44 (0.95 - 2.18) |
| He et al [2024] | A | R7 | Esophageal adenocarcinoma | 4112 | 17159 | 30 | 1.00 (0.90 - 1.11) |
| He et al [2024] | B | R7 | Barrett's esophagus | 946 | 292256 | 32 | 1.14 (0.94 - 1.38) |
| He et al [2024] | B | R7 | Perforation of esophagus | 103 | 292256 | 32 | 1.23 (0.69 - 2.19) |
| He et al [2024] | B | R7 | Oesophageal obstruction | 879 | 292256 | 33 | 0.90 (0.73 - 1.12) |
| He et al [2024] | B | R7 | Esophageal varices | 894 | 295014 | 32 | 1.06 (0.87 - 1.30) |
| He et al [2024] | B | R7 | Esophageal ulcer | 1840 | 292256 | 32 | 1.05 (0.91 - 1.21) |
| He et al [2024] | B | R7 | Benign esophageal tumors | 260 | 342239 | 32 | 0.80 (0.54 - 1.18) |
| He et al [2024] | B | R7 | Esophagitis | 1318 | 292256 | 32 | 1.03 (0.86 - 1.24) |
| He et al [2024] | B | R7 | Gastroesophageal reflux disease | 22867 | 292256 | 33 | 1.00 (0.96 - 1.05) |
| He et al [2024] | B | R7 | Congenital esophageal malformation | 94 | 341501 | 32 | 1.75 (0.97 - 3.17) |
| He et al [2024] | B | R7 | Esophageal adenocarcinoma | 4112 | 17159 | 30 | 1.03 (0.90 - 1.19) |
| Cao et al [2022] | A | R6 | T2DM | 74,124 | 824,006 | 15 | 1.06 (1.00 - 1.13) |
| Cao et al [2022] | B | R6 | T2DM | 74,124 | 824,006 | 20 | 1.09 (0.99 - 1.19) |
| Cao et al [2022] | C | R6 | T2DM | 74,124 | 824,006 | 11 | 1.25 (1.00 - 1.56) |
| Li et al [2022] | C | R5 | Autoimmune thyroid disease | 30,234 | 725,172 | 19 | 1.042 (0.984 - 1.103) |
| Li et al [2022] | B | R5 | Autoimmune thyroid disease | 30,234 | 725,172 | 22 | 1.014 (0.991 - 1.037) |
| Li et al [2022] | A | R5 | Autoimmune thyroid disease | 30,234 | 725,172 | 31 | 0.996 (0.982 - 1.011) |
| Li et al [2022] | C | R5 | Hyperthyroidism | 1,840 | 49,983 | 17 | 1.063 (0.893 - 1.265) |
| Li et al [2022] | B | R5 | Hyperthyroidism | 1,840 | 49,983 | 22 | 1.006 (0.928 - 1.092) |
| Li et al [2022] | A | R5 | Hyperthyroidism | 1,840 | 49,983 | 32 | 0.983 (0.937 - 1.031) |
| Li et al [2022] | C | R5 | Hypothyroidism | 3,440 | 49,983 | 18 | 1.335 (1.167 - 1.526) |
| Li et al [2022] | B | R5 | Hypothyroidism | 3,440 | 49,983 | 20 | 1.037 (0.98 - 1.097) |
| Li et al [2022] | A | R5 | Hypothyroidism | 3,440 | 49,983 | 32 | 1.036 (0.992 - 1.082) |
| Yao et al [2023] | A | R7 | RA | 14,361 | 43,923 | 18 | 1 (0.950 - 1.080) |
| Yao et al [2023] | B | R7 | RA | 14,361 | 43,923 | 18 | 1.02 (0.910 - 1.110) |
| Yao et al [2023] | C | R7 | RA | 14,361 | 43,923 | 7 | 1 (0.570 - 1.700) |
| Yao et al [2023] | A | R7 | SLE | 7,219 | 15,991 | 21 | 1 (0.920 - 1.100) |
| Yao et al [2023] | B | R7 | SLE | 7,219 | 15,991 | 22 | 1.03 (0.900 - 1.190) |
| Yao et al [2023] | C | R7 | SLE | 7,219 | 15,991 | 7 | 0.91 (0.560 - 2.610) |
| Ran et al [2022] | A | R5 | SLE | 5,201 | 9,066 | NA | 0.886 (0.676 - 1.160) |
| Ran et al [2022] | B | R5 | SLE | 5,201 | 9,066 | NA | 1.208 (0.881 - 1.657) |
| Ran et al [2022] | C | R5 | SLE | 5,201 | 9,066 | NA | 0.911 (0.423 - 1.963) |
| Yang et al [2023] | C | R7 | SLE | 5,201 | 9,066 | 12 | 0.89 (0.540 - 1.450) |
| Yang et al [2023] | B | R7 | SLE | 5,201 | 9,066 | 23 | 0.94 (0.750 - 1.180) |
| Yang et al [2023] | A | R7 | SLE | 5,201 | 9,066 | 21 | 1.06 (0.970 - 1.150) |
| Yang et al [2023] | C | R7 | SLE | 4,222 | 8,431 | 3 | 1.06 (0.840 - 1.330) |
| Yang et al [2023] | B | R7 | SLE | 4,222 | 8,431 | 5 | 1.07 (0.900 - 1.280) |
| Yang et al [2023] | A | R7 | SLE | 4,222 | 8,431 | 4 | 0.98 (0.900 - 1.070) |
| Xu et al [2023] | A | R7 | ME/CFS | 2,076 | 460,857 | 20 | 0.99973 (0.999 - 1.000) |
| Xu et al [2023] | B | R7 | ME/CFS | 2,076 | 460,857 | 24 | 0.99994 (0.999 - 1.001) |
| Xu et al [2023] | C | R7 | ME/CFS | 2,076 | 460,857 | 13 | 1 (0.998 - 1.002) |
| Sun et al [2023] | C | R7 | EOMG | 595 | 2,718 | 21 | 0.91 (0.440 - 1.880) |
| Sun et al [2023] | B | R7 | EOMG | 595 | 2,718 | 38 | 1.33 (0.990 - 1.780) |
| Sun et al [2023] | A | R7 | EOMG | 595 | 2,718 | 34 | 1.17 (0.920 - 1.490) |
| Sun et al [2023] | C | R7 | LOMG | 1,278 | 33,652 | 20 | 1.09 (0.670 - 1.780) |
| Sun et al [2023] | B | R7 | LOMG | 1,278 | 33,652 | 37 | 1.06 (0.880 - 1.280) |
| Sun et al [2023] | A | R7 | LOMG | 1,278 | 33,652 | 32 | 1.08 (0.950 - 1.230) |
| Sun et al [2023] | C | R7 | MG | 1,873 | 36,370 | 21 | 1.09 (0.740 - 1.590) |
| Sun et al [2023] | B | R7 | MG | 1,873 | 36,370 | 37 | 1.1 (0.950 - 1.270) |
| Sun et al [2023] | A | R7 | MG | 1,873 | 36,370 | 31 | 1.09 (0.980 - 1.200) |
| Luo et al [2023] | A | R7 | Atopic dermatitis | 53,458 | 324,092 | 26 | 1.04 (0.960 - 1.130) |
| Luo et al [2023] | B | R7 | Atopic dermatitis | 53,458 | 324,092 | 26 | 1.04 (0.960 - 1.130) |
| Luo et al [2023] | C | R7 | Atopic dermatitis | 53,458 | 324,092 | 14 | 1 (0.760 - 1.320) |
| Chalitsios et al [2022] | A | R7 | Psoriasis | 13,229 | 21,543 | NA | 0.930 (0.760 - 1.130) |
| Chalitsios et al [2022] | B | R7 | Psoriasis | 13,229 | 21,543 | NA | 1.1 (0.780 - 1.550) |
| Chalitsios et al [2022] | C | R7 | Psoriasis | 13,229 | 21,543 | NA | 0.910 (0.323 - 2.630) |
| Sun et al [2023] | A | R7 | NMOSD | 215 | 1,244 | 20 | 1.915 (0.647 - 5.667) |
| Sun et al [2023] | B | R7 | NMOSD | 215 | 1,244 | 36 | 1.483 (0.951 - 2.310) |
| Sun et al [2023] | C | R7 | NMOSD | 215 | 1,244 | 31 | 1.309 (0.959 - 1.787) |
| Sun et al [2023] | A | R7 | AQP4+NMOSD | 132 | 1,244 | 20 | 4.958 (1.322 - 18.585) |
| Sun et al [2023] | B | R7 | AQP4+NMOSD | 132 | 1,244 | 35 | 1.656 (0.954 - 2.874) |
| Sun et al [2023] | C | R7 | AQP4+NMOSD | 132 | 1,244 | 31 | 1.354 (0.921 - 1.991) |
| Sun et al [2023] | A | R7 | AQP4-NMOSD | 83 | 1,244 | 20 | 0.565 (0.099 - 3.212) |
| Sun et al [2023] | B | R7 | AQP4-NMOSD | 83 | 1,244 | 33 | 1.416 (0.695 - 2.883) |
| Sun et al [2023] | C | R7 | AQP4-NMOSD | 83 | 1,244 | 29 | 1.296 (0.788 - 2.1383) |
| Quan et al [2023] | C | GenOMICC | Juvenile idiopathic arthritis | 3,553 | 440,646 | NA | 1.517 (1.144 - 2.011) |
| Quan et al [2023] | C | GenOMICC | Primary biliary cholangitis | 2,861 | 8,514 | NA | 1.37 (1.149 - 1.635) |
| Quan et al [2023] | C | GenOMICC | Gout | 2,115 | 67,259 | NA | 1.075 (0.960 - 1.204) |
| Quan et al [2023] | C | GenOMICC | Ankylosing spondylitis | 541 | 74,589 | NA | 1.073 (0.833 - 1.381) |
| Quan et al [2023] | C | GenOMICC | Primary sjogren's syndrome | 496 | 94,550 | NA | 0.995 (0.796 - 1.244) |
| Quan et al [2023] | C | GenOMICC | SLE | 1,311 | 1,783 | NA | 0.732 (0.590 - 0.908) |
| Wang et al [2023] | C | R7 | NMOSD | NA | NA | NA | 2.09 (0.685 - 6.374) |
| Wang et al [2023] | B | R7 | NMOSD | NA | NA | NA | 1.402 (0.89 - 2.209) |
| Wang et al [2023] | A | R7 | NMOSD | NA | NA | NA | 1.124 (0.83 - 1.523) |
| Hao et al [2022] | C | R4 | Gout | NA | NA | 7 | 1.001 (0.999 - 1.003) |
| Hao et al [2022] | B | R4 | Gout | NA | NA | 8 | 1.000 (0.999 - 1.001) |
| Hao et al [2022] | A | R4 | Gout | NA | NA | 7 | 1.000 (0.999 - 1.001) |
| Zou et al [2024] | C | R7 | Celiac disease | 4533 | 10,750 | 4 | 0.938 (0.438 - 2.007) |
| Zou et al [2024] | B | R7 | Celiac disease | 4533 | 10,750 | 6 | 1.082 (0.856 - 1.367) |
| Zou et al [2024] | A | R7 | Celiac disease | 4533 | 10,750 | 7 | 1.115 (1.007 - 1.234) |
| Kong et al [2024] | B | R5 | GBS | 213 | 215,718 | NA | 0.831 (0.596 - 1.159) |
| Wang et al [2024] | C | R5 | T1DM | 18,942 | 501,638 | 7 | 1.164 (0.850 - 1.594) |
| Gao et al [2024] | C | R5 | Zoster | 2,080 | 211,856 | 7 | 0.97 (0.66 - 1.42) |
| Gao et al [2024] | B | R5 | Zoster | 2,080 | 211,856 | 5 | 1.02 (0.86 - 1.20) |
| Gao et al [2024] | A | R5 | Zoster | 2,080 | 211,856 | 8 | 0.93 (0.81 - 1.08) |
| Wang et al [2023] | B | R5 | Male genital cancer | 6,795 | 354,399 | 5 | 1.000 (0.998 - 1.001) |
| Chen et al [2023] | A | R5 | NHL DLBCL | 209 | 174,006 | 7 | 1.765 (1.174 - 2.651) |
| Chen et al [2023] | A | R5 | HL | 369 | 180,756 | 7 | 1.027 (0.680 - 1.551) |
| Chen et al [2023] | A | R5 | FL | 522 | 180,756 | 7 | 1.205 (0.867 - 1.620) |
| Chen et al [2023] | A | R5 | Mature T/NK-cell lymphomas | 150 | 180,756 | 7 | 1.211 (0.750 - 1.955) |
| Chen et al [2023] | A | R5 | Other and unspecified types of NHL | 533 | 180,756 | 7 | 1.187 (0.833 - 1.693) |
| Dong et al [2023] | C | R5 | GBM | 6,183 | 18,169 | 4 | 0.984 (0.676 - 1.434) |
| Dong et al [2023] | A | R5 | GBM | 6,183 | 18,169 | 6 | 1.093 (0.976 - 1.225) |
| Dong et al [2023] | B | R5 | GBM | 6,183 | 18,169 | 5 | 1.202 (1.035 - 1.395) |
| Song et al [2023] | A | R7 | TRAIL | NA | NA | 31 | 1.019 (0.98 - 1.06) |
| Song et al [2023] | A | R7 | TRAIL receptor 2 | NA | NA | 31 | 0.996 (0.956 - 1.037) |
| Song et al [2023] | A | R7 | TNFSF14 | NA | NA | 31 | 1.004 (0.97 - 1.038) |
| Song et al [2023] | A | R7 | TNFR1 | NA | NA | 31 | 0.997 (0.963 - 1.032) |
| Song et al [2023] | A | R7 | Tumor necrosis factor receptor superfamily member 5 levels | NA | NA | 31 | 0.988 (0.956 - 1.021) |
| Song et al [2023] | A | R7 | Tumor necrosis factor receptor superfamily member 6 levels | NA | NA | 31 | 1.014 (0.983 - 1.046) |
| Song et al [2023] | A | R7 | CD40L | NA | NA | 31 | 0.982 (0.952 - 1.013) |
| Li et al [2023] | A | R7 | Bladder cancer | 1,279 | 372,016 | 24 | 1 (0.9995 - 1.0005) |
| Li et al [2023] | B | R7 | Bladder cancer | 1,279 | 372,016 | 27 | 1.0002 (0.9996 - 1.0007) |
| Li et al [2023] | C | R7 | Bladder cancer | 1,279 | 372,016 | 12 | 0.9991 (0.9976 - 1.0006) |
| Li et al [2023] | A | R7 | Brain glioblastoma cancer | 91 | 174,006 | 26 | 0.9407 (0.5909 - 1.4974) |
| Li et al [2023] | B | R7 | Brain glioblastoma cancer | 91 | 174,006 | 30 | 0.8939 (0.4833 - 1.6533) |
| Li et al [2023] | C | R7 | Brain glioblastoma cancer | 91 | 174,006 | 13 | 1.2895 (0.1937 - 8.5843) |
| Li et al [2023] | A | R7 | BC | 122,977 | 105,974 | 23 | 0.9969 (0.9719 - 1.0226) |
| Li et al [2023] | B | R7 | BC | 122,977 | 105,974 | 28 | 1.0013 (0.9600 - 1.0444) |
| Li et al [2023] | C | R7 | BC | 122,977 | 105,974 | 11 | 0.9616 (0.8487 - 1.0895) |
| Li et al [2023] | A | R7 | BC, HER+ | 4,263 | 99,267 | 26 | 1.0924 (1.02 - 1.1699) |
| Li et al [2023] | B | R7 | BC, HER+ | 4,263 | 99,267 | 30 | 1.1096 (1.0019 - 1.229) |
| Li et al [2023] | C | R7 | BC, HER-positive | 4,263 | 99,267 | 13 | 1.3072 (0.9964 - 1.7149) |
| Li et al [2023] | A | R7 | Cervical cancer | 1,889 | 461,044 | 16 | 1.0001 (0.9995 - 1.0006) |
| Li et al [2023] | B | R7 | Cervical cancer | 1,889 | 461,044 | 18 | 1 (0.9992 - 1.0008) |
| Li et al [2023] | C | R7 | Cervical cancer | 1,889 | 461,044 | 10 | 1.0002 (0.9983 - 1.0022) |
| Li et al [2023] | A | R7 | Colon cancer | 1,494 | 461,439 | 14 | 1.0006 (1 - 1.0011) |
| Li et al [2023] | B | R7 | Colon cancer | 1,494 | 461,439 | 17 | 1.0003 (0.9996 - 1.0011) |
| Li et al [2023] | C | R7 | Colon cancer | 1,494 | 461,439 | 10 | 1.0009 (0.9991 - 1.0026) |
| Li et al [2023] | A | R7 | Colorectal cancer | 5,657 | 372,016 | 24 | 1.001 (1.0001 - 1.0018) |
| Li et al [2023] | B | R7 | Colorectal cancer | 5,657 | 372,016 | 27 | 1.001 (0.9997 - 1.0023) |
| Li et al [2023] | C | R7 | Colorectal cancer | 5,657 | 372,016 | 12 | 1.0024 (0.9992 - 1.0055) |
| Li et al [2023] | A | R7 | Corpus uteri cancer | 1,053 | 99,321 | 26 | 0.9093 (0.7701 - 1.0735) |
| Li et al [2023] | B | R7 | Corpus uteri cancer | 1,053 | 99,321 | 30 | 0.8598 (0.6885 - 1.0736) |
| Li et al [2023] | C | R7 | Corpus uteri cancer | 1,053 | 99,321 | 13 | 0.8968 (0.5314 - 1.5134) |
| Li et al [2023] | A | R7 | ER + BC | 69,501 | 105,974 | 23 | 0.998 (0.9709 - 1.0258) |
| Li et al [2023] | B | R7 | ER + BC | 69,501 | 105,974 | 28 | 0.9957 (0.9504 - 1.0431) |
| Li et al [2023] | C | R7 | ER + BC | 69,501 | 105,974 | 11 | 0.9347 (0.8222 - 1.0626) |
| Li et al [2023] | B | R7 | ER- BC | 21,468 | 105,974 | 23 | 1.0221 (0.9592 - 1.089) |
| Li et al [2023] | C | R7 | ER- BC | 21,468 | 105,974 | 28 | 1.0001 (0.9989 - 1.0012) |
| Li et al [2023] | A | R7 | ER- BC | 21,468 | 105,974 | 11 | 1.0032 (0.9591 - 1.0494) |
| Li et al [2023] | A | R7 | Eye and adnexa cancer | 161 | 174,006 | 26 | 1.1019 (0.7935 - 1.5302) |
| Li et al [2023] | B | R7 | Eye and adnexa cancer | 161 | 174,006 | 30 | 1.1311 (0.7125 - 1.7955) |
| Li et al [2023] | C | R7 | Eye and adnexa cancer | 161 | 174,006 | 13 | 1.6482 (0.383 - 7.0933) |
| Li et al [2023] | A | R7 | Haemotological cancer | 4,552 | 372,016 | 24 | 1.0003 (0.9996 - 1.0011) |
| Li et al [2023] | B | R7 | Haemotological cancer | 4,552 | 372,016 | 27 | 1.0006 (0.9992 - 1.0019) |
| Li et al [2023] | C | R7 | Haemotological cancer | 4,552 | 372,016 | 12 | 0.9894 (0.8241 - 1.1879) |
| Li et al [2023] | A | R7 | Head and neck cancer | 1,106 | 372,016 | 23 | 0.9998 (0.9994 - 1.0003) |
| Li et al [2023] | B | R7 | Head and neck cancer | 1,106 | 372,016 | 26 | 0.9998 (0.9991 - 1.0004) |
| Li et al [2023] | C | R7 | Head and neck cancer | 1,106 | 372,016 | 12 | 0.9986 (0.9972 - 1) |
| Li et al [2023] | A | R7 | Heart, mediastinum and pleura cancer | 102 | 174,006 | 26 | 0.8622 (0.5739 - 1.2952) |
| Li et al [2023] | B | R7 | Heart, mediastinum and pleura cancer | 102 | 174,006 | 29 | 0.868 (0.4861 - 1.5499) |
| Li et al [2023] | C | R7 | Heart, mediastinum and pleura cancer | 102 | 174,006 | 13 | 0.7427 (0.1483 - 3.7192) |
| Li et al [2023] | C | R7 | Kidney, except renal pelvis cancer | 971 | 174,977 | 26 | 0.8063 (0.4607 - 1.4111) |
| Li et al [2023] | B | R7 | Kidney, except renal pelvis cancer | 971 | 174,977 | 30 | 0.9006 (0.7319 - 1.1082) |
| Li et al [2023] | A | R7 | Kidney, except renal pelvis cancer | 971 | 174,977 | 13 | 0.9852 (0.8557 - 1.1343) |
| Li et al [2023] | A | R7 | Laryngeal cancer | 273 | 372,016 | 23 | 1.0001 (0.9999 - 1.0003) |
| Li et al [2023] | B | R7 | Laryngeal cancer | 273 | 372,016 | 23 | 1 (0.9998 - 1.0005) |
| Li et al [2023] | C | R7 | Laryngeal cancer | 273 | 372,016 | 12 | 0.9999 (0.9992 - 1.0006) |
| Li et al [2023] | B | R7 | Lip, oral cavity and pharynx cancer | 126 | 174,006 | 26 | 0.8214 (0.4889 - 1.3802) |
| Li et al [2023] | A | R7 | Lip, oral cavity and pharynx cancer | 126 | 174,006 | 30 | 0.7843 (0.5425 - 1.134) |
| Li et al [2023] | C | R7 | Lip, oral cavity and pharynx cancer | 126 | 174,006 | 13 | 0.2821 (0.0527 - 1.5107) |
| Li et al [2023] | A | R7 | Liver & bile duct cancer | 350 | 372,016 | 23 | 0.9999 (0.9997 - 1.0001) |
| Li et al [2023] | B | R7 | Liver & bile duct cancer | 350 | 372,016 | 23 | 0.9999 (0.9996 - 1.0002) |
| Li et al [2023] | C | R7 | Liver & bile duct cancer | 350 | 372,016 | 12 | 1.0001 (0.9993 - 1.0009) |
| Li et al [2023] | A | R7 | Lung cancer | 2,671 | 372,016 | 24 | 0.9996 (0.9989 - 1.0003) |
| Li et al [2023] | B | R7 | Lung cancer | 2,671 | 372,016 | 27 | 0.9995 (0.9984 - 1.0006) |
| Li et al [2023] | C | R7 | Lung cancer | 2,671 | 372,016 | 12 | 0.9981 (0.9958 - 1.0004) |
| Li et al [2023] | A | R7 | Melanoma skin cancer | 3,751 | 372,016 | 24 | 0.9995 (0.9988 - 1.0003) |
| Li et al [2023] | B | R7 | Melanoma skin cancer | 3,751 | 372,016 | 27 | 0.9993 (0.9982 - 1.0005) |
| Li et al [2023] | C | R7 | Melanoma skin cancer | 3,751 | 372,016 | 12 | 1.0008 (0.997 - 1.0046) |
| Li et al [2023] | A | R7 | Meninges cancer | 640 | 174,006 | 26 | 0.9926 (0.8273 - 1.1908) |
| Li et al [2023] | B | R7 | Meninges cancer | 640 | 174,006 | 30 | 1.0748 (0.8224 - 1.4047) |
| Li et al [2023] | C | R7 | Meninges cancer | 640 | 174,006 | 13 | 1.7574 (0.8541 - 3.6161) |
| Li et al [2023] | B | R7 | Non-small cell lung cancer | 1,627 | 174,006 | 26 | 1.0065 (0.8558 - 1.1836) |
| Li et al [2023] | C | R7 | Non-small cell lung cancer | 1,627 | 174,006 | 30 | 1.0486 (0.6681 - 1.6459) |
| Li et al [2023] | A | R7 | Non-small cell lung cancer | 1,627 | 174,006 | 13 | 1.0059 (0.9033 - 1.1202) |
| Li et al [2023] | A | R7 | Oesophageal cancer | NA | NA | 23 | 1.0004 (1.0001 - 1.0007) |
| Li et al [2023] | B | R7 | Oesophageal cancer | NA | NA | 25 | 1.0005 (1 - 1.0009) |
| Li et al [2023] | C | R7 | Oesophageal cancer | NA | NA | 12 | 1.0304 (0.6441 - 1.6484) |
| Li et al [2023] | A | R7 | Ovarian cancer | 1,218 | 198,523 | 24 | 1.0007 (0.9999 - 1.0016) |
| Li et al [2023] | B | R7 | Ovarian cancer | 1,218 | 198,523 | 27 | 1.001 (0.9997 - 1.0023) |
| Li et al [2023] | C | R7 | Ovarian cancer | 1,218 | 198,523 | 12 | 1.0021 (0.9987 - 1.0054) |
| Li et al [2023] | A | R7 | Pancreas cancer | 3,022 | 174,006 | 26 | 1.0279 (0.8660 - 1.22) |
| Li et al [2023] | B | R7 | Pancreas cancer | 3,022 | 174,006 | 30 | 1.0952 (0.8605 - 1.394) |
| Li et al [2023] | C | R7 | Pancreas cancer | 3,022 | 174,006 | 13 | 1.848 (0.8519 - 4.0084) |
| Li et al [2023] | A | R7 | Prostate cancer | 79,148 | 61,106 | 24 | 1.0103 (0.9735 - 1.0486) |
| Li et al [2023] | B | R7 | Prostate cancer | 79,148 | 61,106 | 27 | 0.9926 (0.9323 - 1.0568) |
| Li et al [2023] | C | R7 | Prostate cancer | 79,148 | 61,106 | 12 | 0.9889 (0.8988 - 1.088) |
| Li et al [2023] | A | R7 | Rectum cancer | 1,470 | 461,540 | 14 | 1.0002 (0.9995 - 1.0008) |
| Li et al [2023] | B | R7 | Rectum cancer | 1,470 | 461,540 | 17 | 1.0001 (0.9991 - 1.001) |
| Li et al [2023] | C | R7 | Rectum cancer | 1,470 | 461,540 | 10 | 1.0008 (0.9988 - 1.0028) |
| Li et al [2023] | A | R7 | Small cell lung cancer | 179 | 174,006 | 26 | 0.7895 (0.5548 - 1.1234) |
| Li et al [2023] | B | R7 | Small cell lung cancer | 179 | 174,006 | 30 | 0.7513 (0.4842 - 1.1658) |
| Li et al [2023] | C | R7 | Small cell lung cancer | 179 | 174,006 | 13 | 0.5593 (0.1622 - 1.9288) |
| Li et al [2023] | A | R7 | Small intestine/small bowel cancer | 156 | 337,003 | 26 | 1 (0.9999 - 1.0002) |
| Li et al [2023] | B | R7 | Small intestine/small bowel cancer | 156 | 337,003 | 30 | 1 (0.9999 - 1.0003) |
| Li et al [2023] | C | R7 | Small intestine/small bowel cancer | 156 | 337,003 | 13 | 1.0001 (0.9994 - 1.0007) |
| Li et al [2023] | A | R7 | Stomach cancer | 633 | 174,006 | 26 | 1.2394 (1.0173 - 1.5099) |
| Li et al [2023] | B | R7 | Stomach cancer | 633 | 174,006 | 30 | 1.3043 (1.0028 - 1.6964) |
| Li et al [2023] | C | R7 | Stomach cancer | 633 | 174,006 | 13 | 2.8563 (1.4747 - 5.5323) |
| Li et al [2023] | A | R7 | Testis cancer | 199 | 74,685 | 26 | 0.8175 (0.5995 - 1.1149) |
| Li et al [2023] | B | R7 | Testis cancer | 199 | 74,685 | 30 | 0.852 (0.5449 - 1.3323) |
| Li et al [2023] | C | R7 | Testis cancer | 199 | 74,685 | 13 | 0.463 (0.1440 - 1.4889) |
| Li et al [2023] | A | R7 | Thyroid cancer | 989 | 174,006 | 26 | 0.9245 (0.8073 - 1.0587) |
| Li et al [2023] | C | R7 | Thyroid cancer | 989 | 174,006 | 30 | 0.5896 (0.3451 - 1.0072) |
| Li et al [2023] | B | R7 | Thyroid cancer | 989 | 174,006 | 13 | 0.9437 (0.7797 - 1.1421) |
| Li et al [2023] | A | R7 | Vulva cancer | 125 | 99,321 | 26 | 0.8942 (0.6114 - 1.3077) |
| Li et al [2023] | B | R7 | Vulva cancer | 125 | 99,321 | 30 | 1.0582 (0.6174 - 1.8138) |
| Li et al [2023] | C | R7 | Vulva cancer | 125 | 99,321 | 13 | 0.8751 (0.1736 - 4.4112) |
| Zhao et al [2023] | C | R7 | BC | 133,384 | 113,789 | 16 | 0.99 (0.86 - 1.114) |
| Zhao et al [2023] | B | R7 | BC | 133,384 | 113,789 | 37 | 1 (0.96 - 1.05) |
| Zhao et al [2023] | A | R7 | BC | 133,384 | 113,789 | 36 | 1.01 (0.98 - 1.04) |
| Zhao et al [2023] | C | R7 | EC | 22,406 | 40,941 | 16 | 1.26 (0.97 - 1.64) |
| Zhao et al [2023] | B | R7 | EC | 22,406 | 40,941 | 37 | 1.09 (1.01 - 1.18) |
| Zhao et al [2023] | A | R7 | EC | 22,406 | 40,941 | 36 | 1.06 (1 - 1.11) |
| Zhao et al [2023] | C | R7 | EOC | 12,906 | 108,979 | 16 | 1.19 (0.95 - 1.48) |
| Zhao et al [2023] | B | R7 | EOC | 12,906 | 108,979 | 36 | 1.01 (0.92 - 1.1) |
| Zhao et al [2023] | A | R7 | EOC | 12,906 | 108,979 | 35 | 1.06 (0.92 - 1.06) |
| Jin et al [2023] | A | R7 | Colon cancer | 1,803 | 216,989 | NA | 1.0459 (0.9355 - 1.1694) |
| Jin et al [2023] | B | R7 | Colon cancer | 1,803 | 216,989 | NA | 1.0920 (0.9193 - 1.2971) |
| Jin et al [2023] | C | R7 | Colon cancer | 1,803 | 216,989 | NA | 1.0738 (0.6307 - 1.8282) |
| Jin et al [2023] | A | R7 | Colorectal cancer | 3,022 | 215,770 | NA | 1.0092 (0.9234 - 1.1030) |
| Jin et al [2023] | B | R7 | Colorectal cancer | 3,022 | 215,770 | NA | 1.0802 (0.9520 - 1.2256) |
| Jin et al [2023] | C | R7 | Colorectal cancer | 3,022 | 215,770 | NA | 1.0157 (0.7066 - 1.4600) |
| Jin et al [2023] | A | R7 | Esophageal cancer | 232 | 218,560 | NA | 0.9975 (0.7184 - 1.3848) |
| Jin et al [2023] | B | R7 | Esophageal cancer | 232 | 218,560 | NA | 0.9316 (0.6207 - 1.3982) |
| Jin et al [2023] | C | R7 | Esophageal cancer | 232 | 218,560 | NA | 1.1435 (0.3207 - 4.0772) |
| Jin et al [2023] | A | R7 | Stomach cancer | 633 | 218,159 | NA | 1.2301 (1.0075 - 1.5020) |
| Jin et al [2023] | B | R7 | Stomach cancer | 633 | 218,159 | NA | 1.3177 (1.0166 - 1.7080) |
| Jin et al [2023] | C | R7 | Stomach cancer | 633 | 218,159 | NA | 1.4896 (0.5960 - 3.7232) |
| Xu et al [2023] | C | R5 | Thyroid cancer | 649 | 431 | 9 | 2.826 (0.842 - 9.483) |
| Xu et al [2023] | B | R5 | Thyroid cancer | 649 | 431 | 11 | 1.630 (1.050 - 2.529) |
| Xu et al [2023] | A | R5 | Thyroid cancer | 649 | 431 | 12 | 1.061 (0.575 - 1.956) |
| Wu et al [2022] | C | R4 | Endometrial cancer | 12,906 | 108,979 | 13 | 1.165 (1.012 - 1.340) |
| Wu et al [2022] | B | R4 | Endometrial cancer | 12,906 | 108,979 | 7 | 1.145 (1.002 - 1.309) |
| Wu et al [2022] | A | R4 | Endometrial cancer | 12,906 | 108,979 | 7 | 1.075 (1.006 - 1.148) |
| Wang et al [2024] | C | R5 | Laryngeal cancer | 4,792 | 1,054,664 | 7 | 0.24 (0.05 - 1.26) |
| Wang et al [2024] | B | R5 | Laryngeal cancer | 4,792 | 1,054,664 | 5 | 0.51 (0.27 - 0.95) |
| Wang et al [2024] | A | R5 | Laryngeal cancer | 4,792 | 1,054,664 | 7 | 0.62 (0.43 - 0.90) |
| Han et al [2024] | C | R7 | HNC | 2,131 | 287,137 | 14 | 0.52 (0.35 - 0.78) |
| Han et al [2024] | B | R7 | HNC | 2,131 | 287,137 | 32 | 0.99 (0.84 - 1.17) |
| Han et al [2024] | A | R7 | HNC | 2,131 | 287,137 | 27 | 0.95 (0.84 - 1.07) |
| Song et al [2024] | A | R7 | Lung cancer | 29,266 | 56,450 | 10 | 1.01 (0.78 - 1.30) |
| Song et al [2024] | A | R7 | Lung adenocarcinoma | 11,273 | 55,483 | 11 | 0.91 (0.62 - 1.33) |
| Song et al [2024] | A | R7 | Lung squamous cell carcinoma | 7,426 | 55,627 | 11 | 0.84 (0.57 - 1.24) |
| Song et al [2024] | A | R7 | Small-cell lung cancer | 2,664 | 21,444 | 11 | 1.29 (0.85 - 1.95) |
| Lu et al [2023] | A | R7 | Sepsis | 10,154 | 454,764 | 30 | 1.000 (0.956 - 1.046) |
| Lu et al [2023] | B | R7 | Sepsis | 10,154 | 454,764 | 36 | 0.976 (0.92 - 1.036) |
| Lu et al [2023] | C | R7 | Sepsis | 10,154 | 454,764 | 17 | 0.923 (0.796 - 1.071) |
| Liu et al [2023] | C | R7 | HBV | 1,394 | 211,059 | 53 | 0.94 (0.82 - 1.08) |
| Liu et al [2023] | B | R7 | HBV | 1,394 | 211,059 | 53 | 1.00 (0.90 - 1.11) |
| Liu et al [2023] | A | R7 | HBV | 1,394 | 211,059 | 45 | 1.02 (0.95 - 1.09) |
| Song et al [2023] | C | R5 | Periodontitis | 12,289 | 22,326 | 4 | 1.04 (0.88 - 1.21) |
| Song et al [2023] | B | R5 | Periodontitis | 12,289 | 22,326 | 1 | 0.97 (0.78 - 1.2) |
| Song et al [2023] | A | R5 | Periodontitis | 12,289 | 22,326 | 6 | 1.01 (0.92 - 1.11) |
| Shi et al [2022] | C | R4 | Miscarriage | 79,047 | 168,133 | 7 | 0.9981 (0.9923 - 1.004) |
| Shi et al [2022] | C | R4 | Miscarriage | 79,048 | 168,134 | 7 | 0.9981 (0.9872 - 1.0091) |
| Chang et al [2023] | C | R5 | Acute renal failure | 212,841 | 215,224 | 7 | 0.99987959 (0.99830243 - 1.00145924) |
| Chang et al [2023] | A | R5 | Acute renal failure | 212,843 | 215,226 | 8 | 1.00027626 (0.99964633 - 1.00090658) |
| Chang et al [2023] | B | R5 | Acute renal failure | 212,842 | 215,225 | 5 | 1.0778966 (0.9225454 - 1.259408) |
| Chang et al [2023] | A | R5 | ATIN | 201,028 | 212,244 | 8 | 0.978838 (0.932636 - 1.027329) |
| Chang et al [2023] | B | R5 | ATIN | 201,028 | 212,244 | 5 | 0.9712434 (0.9028902 - 1.044771) |
| Chang et al [2023] | C | R5 | ATIN | 201,028 | 212,244 | 7 | 0.9540028 (0.819082 - 1.111148) |
| Chang et al [2023] | A | R5 | Benign neoplasm: Adrenal gland | 180,264 | 180,989 | 8 | 1.0866734 (0.905027 - 1.304778) |
| Chang et al [2023] | B | R5 | Benign neoplasm: Adrenal gland | 180,264 | 180,989 | 5 | 1.036893 (0.7715903 - 1.393418) |
| Chang et al [2023] | C | R5 | Benign neoplasm: Adrenal gland | 180,264 | 180,989 | 7 | 1.0134094 (0.56954264 - 1.803199) |
| Chang et al [2023] | A | R5 | Benign neoplasm: Bladder | 180,709 | 180,818 | 8 | 0.8899644 (0.5618528 - 1.409687) |
| Chang et al [2023] | B | R5 | Benign neoplasm: Bladder | 180,709 | 180,818 | 5 | 0.9113894 (0.42546 - 1.952312) |
| Chang et al [2023] | C | R5 | Benign neoplasm: Bladder | 180,709 | 180,818 | 7 | 3.012806 (0.7119013 - 12.75037) |
| Chang et al [2023] | A | R5 | Benign neoplasm: Kidney | 180,506 | 180,887 | 8 | 0.9157382 (0.7118862 - 1.177964) |
| Chang et al [2023] | B | R5 | Benign neoplasm: Kidney | 180,506 | 180,887 | 5 | 0.9159844 (0.6262499 - 1.339764) |
| Chang et al [2023] | C | R5 | Benign neoplasm: Kidney | 180,506 | 180,887 | 7 | 0.9372586 (0.4245568 - 2.069108) |
| Chang et al [2023] | A | R5 | Benign neoplasm: prostate | 78,213 | 78,310 | 8 | 0.7027564 (0.4337569 - 1.138579) |
| Chang et al [2023] | B | R5 | Benign neoplasm: prostate | 78,213 | 78,310 | 5 | 0.8740234 (0.421013 - 1.814474) |
| Chang et al [2023] | C | R5 | Benign neoplasm: prostate | 78,213 | 78,310 | 7 | 1.780751 (0.3878784 - 8.17543) |
| Chang et al [2023] | B | R5 | Bladder neck obstruction | NA | NA | 7 | 1.000372 (0.9992983 - 1.001446) |
| Chang et al [2023] | A | R5 | Calculus of kidney and ureter | 213,445 | 218,414 | 7 | 1.0004365 (0.9998973 - 1.000976) |
| Chang et al [2023] | B | R5 | Calculus of kidney and ureter | 213,445 | 218,414 | 8 | 1.00010561 (0.99930217 - 1.0009097) |
| Chang et al [2023] | C | R5 | Calculus of kidney and ureter | 213,445 | 218,414 | 5 | 0.99958474 (0.99828277 - 1.0008884) |
| Chang et al [2023] | A | R5 | Calculus of lower urinary tract | 213,445 | 214,067 | 7 | 1.280692 (1.028007 - 1.595487) |
| Chang et al [2023] | B | R5 | Calculus of lower urinary tract | 213,445 | 214,067 | 8 | 1.412105 (0.9472167 - 2.105159) |
| Chang et al [2023] | C | R5 | Calculus of lower urinary tract | 213,445 | 214,067 | 5 | 1.0466783 (0.40679586 - 2.693084) |
| Chang et al [2023] | A | R5 | CKD | 212,841 | 216,743 | 7 | 0.9787433 (0.9018629 - 1.062178) |
| Chang et al [2023] | B | R5 | CKD | 212,841 | 216,743 | 8 | 0.9748949 (0.88928442 - 1.06874701) |
| Chang et al [2023] | C | R5 | CKD | 212,841 | 216,743 | 5 | 1.08264196 (0.93979338 - 1.24720352) |
| Chang et al [2023] | A | R5 | CTIN | 201,028 | 201,648 | 7 | 0.9773955 (0.7593061 - 1.258125) |
| Chang et al [2023] | B | R5 | CTIN | 201,028 | 201,648 | 8 | 1.0153331 (0.7532717 - 1.368565) |
| Chang et al [2023] | C | R5 | CTIN | 201,028 | 201,648 | 5 | 1.426039 (0.72495901 - 2.805107) |
| Chang et al [2023] | A | R5 | Cystitis | 195,140 | 203,221 | 7 | 0.99957634 (0.99893905 - 1.00021404) |
| Chang et al [2023] | B | R5 | Cystitis | 195,140 | 203,221 | 8 | 0.99909959 (0.99822069 - 0.99997925) |
| Chang et al [2023] | C | R5 | Cystitis | 195,140 | 203,221 | 5 | 0.99767102 (0.99604125 - 0.99930346) |
| Chang et al [2023] | A | R5 | ED | 94,024 | 95,178 | 7 | 1.05403449 (0.9699633 - 1.14539252) |
| Chang et al [2023] | B | R5 | ED | 94,024 | 95,178 | 8 | 1.062475 (0.94110606 - 1.19949618) |
| Chang et al [2023] | C | R5 | ED | 94,024 | 95,178 | 5 | 1.12143304 (0.91438355 - 1.37536602) |
| Chang et al [2023] | A | R5 | Genital infection | NA | NA | 7 | 0.96146284 (0.87616704 - 1.05506228) |
| Chang et al [2023] | B | R5 | Genital infection | NA | NA | 8 | 1.03807122 (0.88765528 - 1.2139756) |
| Chang et al [2023] | C | R5 | Genital infection | NA | NA | 5 | 1.0457597 (0.77631253 - 1.40872818) |
| Chang et al [2023] | C | R5 | Genitourinary benign neoplasm | NA | NA | 5 | 1.13838864 (0.7436988 - 1.7425451) |
| Chang et al [2023] | C | R5 | Genitourinary malignant neoplasm | NA | NA | 8 | 0.99962293 (0.99876404 - 1.00048256) |
| Chang et al [2023] | A | R5 | Genitourinary obstruction | NA | NA | 8 | 1.00013586 (0.99963818 - 1.00063379) |
| Chang et al [2023] | B | R5 | Genitourinary obstruction | NA | NA | 5 | 1.00012315 (0.999446 - 1.00080076) |
| Chang et al [2023] | C | R5 | Genitourinary obstruction | NA | NA | 7 | 1.00008441 (0.99903603 - 1.00113389) |
| Chang et al [2023] | A | R5 | Hydrocele | 72,799 | 75,433 | 8 | 1.0233477 (0.927033 - 1.129669) |
| Chang et al [2023] | B | R5 | Hydrocele | 72,799 | 75,433 | 5 | 1.035115 (0.8916958 - 1.201602) |
| Chang et al [2023] | C | R5 | Hydrocele | 72,799 | 75,433 | 7 | 0.9473759 (0.6633629 - 1.352987) |
| Chang et al [2023] | A | R5 | Hydronephrosis | 201,028 | 202,482 | 8 | 0.9563259 (0.7798362 - 1.172758) |
| Chang et al [2023] | B | R5 | Hydronephrosis | 201,028 | 202,482 | 5 | 0.988035 (0.8122136 - 1.201917) |
| Chang et al [2023] | C | R5 | Hydronephrosis | 201,028 | 202,482 | 7 | 0.8404347 (0.5595266 - 1.262372) |
| Chang et al [2023] | A | R5 | Hyperplasia of prostate | 72,799 | 85,917 | 8 | 1.00025769 (0.99926762 - 1.00124874) |
| Chang et al [2023] | B | R5 | Hyperplasia of prostate | 72,799 | 85,917 | 5 | 1.00054413 (0.998987 - 1.00210369) |
| Chang et al [2023] | C | R5 | Hyperplasia of prostate | 72,799 | 85,917 | 7 | 1.00154189 (0.99944316 - 1.00364503) |
| Chang et al [2023] | A | R5 | Hypertensive Renal Disease | 162,837 | 163,305 | 8 | 0.9161877 (0.7232978 - 1.160518) |
| Chang et al [2023] | B | R5 | Hypertensive Renal Disease | 162,837 | 163,305 | 5 | 1.02344 (0.7244847 - 1.445759) |
| Chang et al [2023] | C | R5 | Hypertensive Renal Disease | 162,837 | 163,305 | 7 | 0.8359664 (0.38450563 - 1.817502) |
| Chang et al [2023] | C | R5 | Kidney infection | NA | NA | 7 | 0.998852 (0.9965797 - 1.00113) |
| Chang et al [2023] | A | R5 | Malignant neoplasm of bladder | 174,006 | 175,121 | 7 | 0.99943805 (0.99883935 - 1.00003711) |
| Chang et al [2023] | B | R5 | Malignant neoplasm of bladder | 174,006 | 175,121 | 8 | 0.99937165 (0.99846723 - 1.00027689) |
| Chang et al [2023] | C | R5 | Malignant neoplasm of bladder | 174,006 | 175,121 | 5 | 0.99938619 (0.99770614 - 1.00106907) |
| Chang et al [2023] | A | R5 | Malignant neoplasm of kidney | 174,006 | 174,977 | 7 | 1.00026618 (0.99979285 - 1.00073973) |
| Chang et al [2023] | B | R5 | Malignant neoplasm of kidney | 174,006 | 174,977 | 8 | 1.00062553 (0.99938964 - 1.00186295) |
| Chang et al [2023] | C | R5 | Malignant neoplasm of kidney | 174,006 | 174,977 | 5 | 1.00019763 (0.99862415 - 1.00177358) |
| Chang et al [2023] | A | R5 | Malignant neoplasm of prostate | 74,685 | 80,996 | 7 | 0.99968551 (0.99878459 - 1.00058723) |
| Chang et al [2023] | B | R5 | Malignant neoplasm of prostate | 74,685 | 80,996 | 8 | 0.99955004 (0.9982111 - 1.00089077) |
| Chang et al [2023] | C | R5 | Malignant neoplasm of prostate | 74,685 | 80,996 | 5 | 0.99937515 (0.99738878 - 1.00136548) |
| Chang et al [2023] | A | R5 | Malignant neoplasm of testis | 74,685 | 74,884 | 8 | 0.8990345 (0.634504 - 1.27385) |
| Chang et al [2023] | B | R5 | Malignant neoplasm of testis | 74,685 | 74,884 | 5 | 0.7172938 (0.424112 - 1.213147) |
| Chang et al [2023] | A | R5 | Neuromuscular dysfuntion of bladder | 195,140 | 196,427 | 8 | 0.9999652 (0.99949972 - 1.0004309) |
| Chang et al [2023] | B | R5 | Neuromuscular dysfuntion of bladder | 195,140 | 196,427 | 5 | 1.00025368 (0.99955024 - 1.00095761) |
| Chang et al [2023] | C | R5 | Neuromuscular dysfuntion of bladder | 195,140 | 196,427 | 7 | 1.00065226 (0.99923999 - 1.00206652) |
| Chang et al [2023] | A | R5 | Orchitis and epididymitis | 72,799 | 74,096 | 8 | 0.9940815 (0.8536672 - 1.157592) |
| Chang et al [2023] | B | R5 | Orchitis and epididymitis | 72,799 | 74,096 | 5 | 1.077478 (0.8584988 - 1.352312) |
| Chang et al [2023] | C | R5 | Orchitis and epididymitis | 72,799 | 74,096 | 7 | 1.283089 (0.8286262 - 1.986802) |
| Chang et al [2023] | A | R5 | Prostatitis | NA | NA | 8 | 0.9426328 (0.8383443 - 1.059895) |
| Chang et al [2023] | B | R5 | Prostatitis | NA | NA | 5 | 1.003696 (0.8087203 - 1.245678) |
| Chang et al [2023] | C | R5 | Prostatitis | NA | NA | 7 | 0.8758735 (0.5829694 - 1.315943) |
| Chang et al [2023] | A | R5 | Renal failure | 212,841 | 218,792 | 8 | 0.9896164 (0.9257575 - 1.05788) |
| Chang et al [2023] | B | R5 | Renal failure | 212,841 | 218,792 | 5 | 1.00037036 (0.99950495 - 1.00123653) |
| Chang et al [2023] | C | R5 | Renal failure | 212,841 | 218,792 | 7 | 0.99987839 (0.99830128 - 1.001458) |
| Chang et al [2023] | A | R5 | TIN | 201,028 | 218,792 | 8 | 0.97878694 (0.93339316 - 1.02638834) |
| Chang et al [2023] | B | R5 | TIN | 201,028 | 218,792 | 5 | 0.97272353 (0.83827952 - 1.12872978) |
| Chang et al [2023] | C | R5 | TIN | 201,028 | 218,792 | 7 | 0.97272353 (0.83827952 - 1.12872978) |
| Chang et al [2023] | A | R5 | Retention of urine | 202,910 | 207,909 | 8 | 0.99986216 (0.99909572 - 1.0006292) |
| Chang et al [2023] | B | R5 | Retention of urine | 202,910 | 207,909 | 5 | 0.99969077 (0.99863744 - 1.00074522) |
| Chang et al [2023] | C | R5 | Retention of urine | 202,910 | 207,909 | 7 | 1.00030794 (0.99838666 - 1.00223291) |
| Chang et al [2023] | A | R5 | Sexual dysfunction | 213,826 | 213,986 | 8 | 1.08081414 (1.00097538 - 1.16702092) |
| Chang et al [2023] | B | R5 | Sexual dysfunction | 213,826 | 213,986 | 5 | 1.10660053 (0.98881378 - 1.23841795) |
| Chang et al [2023] | C | R5 | Sexual dysfunction | 213,826 | 213,986 | 7 | 1.13320164 (0.93312916 - 1.37617171) |
| Chang et al [2023] | A | R5 | Testicular dysfunction | 92,895 | 93,180 | 8 | 1.26370288 (1.02239036 - 1.56197186) |
| Chang et al [2023] | B | R5 | Testicular dysfunction | 92,895 | 93,180 | 5 | 1.49538053 (1.08661982 - 2.05790736) |
| Chang et al [2023] | C | R5 | Testicular dysfunction | 92,895 | 93,180 | 7 | 1.53677171 (0.77075491 - 3.06409633) |
| Chang et al [2023] | A | R5 | Torsion of testis | 72,799 | 73,108 | 8 | 0.9087109 (0.6211761 - 1.329342) |
| Chang et al [2023] | B | R5 | Torsion of testis | 72,799 | 73,108 | 5 | 0.7429304 (0.4862988 - 1.134993) |
| Chang et al [2023] | C | R5 | Torsion of testis | 72,799 | 73,108 | 7 | 0.5379912 (0.2207114 - 1.311371) |
| Chang et al [2023] | A | R5 | T1DKD | 183,185 | 184,148 | 8 | 0.8959585 (0.7618109 - 1.053728) |
| Chang et al [2023] | B | R5 | T1DKD | 183,185 | 184,148 | 5 | 1.0067973 (0.7891497 - 1.284472) |
| Chang et al [2023] | C | R5 | T1DKD | 183,185 | 184,148 | 7 | 1.015426 (0.5690534 - 1.811938) |
| Chang et al [2023] | A | R5 | T2DKD | 183,185 | 184,481 | 8 | 0.9829043 (0.8534989 - 1.13193) |
| Chang et al [2023] | B | R5 | T2DKD | 183,185 | 184,481 | 5 | 1.027716 (0.7942896 - 1.329741) |
| Chang et al [2023] | C | R5 | T2DKD | 183,185 | 184,481 | 7 | 0.9786645 (0.6127476 - 1.563097) |
| Chang et al [2023] | A | R5 | Urinary tract infection | NA | NA | 8 | 0.99972019 (0.99915393 - 1.00028677) |
| Chang et al [2023] | B | R5 | Urinary tract infection | NA | NA | 5 | 0.99918346 (0.99841337 - 0.99995414) |
| Chang et al [2023] | C | R5 | Urinary tract infection | NA | NA | 7 | 0.99807243 (0.99674769 - 0.99939893) |
| Chang et al [2023] | A | R5 | Urolithiasis | 213,445 | 218,792 | 8 | 1.00039348 (0.99988024 - 1.00090698) |
| Chang et al [2023] | B | R5 | Urolithiasis | 213,445 | 218,792 | 5 | 1.00011751 (0.99936609 - 1.00086949) |
| Chang et al [2023] | C | R5 | Urolithiasis | 213,445 | 218,792 | 7 | 0.99933343 (0.99825451 - 1.00041351) |
| Chang et al [2023] | A | R5 | Urological benign neoplasm | NA | NA | 5 | 0.9824204 (0.85815975 - 1.12467387) |
| Chang et al [2023] | B | R5 | Urological benign neoplasm | NA | NA | 7 | 0.97285796 (0.78592616 - 1.2042513) |
| Chang et al [2023] | A | R5 | Urological malignant neoplasm | NA | NA | 7 | 0.99995778 (0.9996589 - 1.00025675) |
| Chang et al [2023] | B | R5 | Urological malignant neoplasm | NA | NA | 8 | 0.99999484 (0.99946933 - 1.00052061) |
| Zhang et al [2022] | C | R4 | ED | 6,175 | 217,630 | 6 | 1.235 (1.044 - 1.462) |
| Si et al [2023] | C | R7 | PCOS | 10,074 | 103,164 | 13 | 1.041 (0.657 - 1.649) |
| Si et al [2023] | B | R7 | PCOS | 10,074 | 103,164 | 30 | 0.995 (0.85 - 1.164) |
| Si et al [2023] | A | R7 | PCOS | 10,074 | 103,164 | 25 | 0.944 (0.843 - 1.058) |
| Liu et al [2024] | B | R5 | Male infertility | 680 | 72,799 | 5 | 0.86 (0.65 - 1.15) |
| Liu et al [2024] | A | R5 | Male infertility | 680 | 72,799 | 8 | 0.93 (0.77 - 1.12) |
| Lin et al [2024] | A | R5 | CKD | 12,385 | 104,780 | 6 | 1.03 (0.96 - 1.10) |
| Ma et al [2024] | A | R7 | ED | 6,175 | 217,630 | 21 | 1.015 (0.957 - 1.078) |
| Ma et al [2024] | B | R7 | ED | 6,175 | 217,630 | 25 | 1.003 (0.922 - 1.093) |
| Ma et al [2024] | C | R7 | ED | 6,175 | 217,630 | 11 | 1.005 (0.781 - 1.293) |
| Ma et al [2024] | A | R7 | T2DM | 48,236 | 250,671 | NA | 1.002 (0.850 - 1.180) |
| Ma et al [2024] | B | R7 | T2DM | 48,236 | 250,671 | NA | 1.011 (0.972 - 1.053) |
| Ma et al [2024] | C | R7 | T2DM | 48,236 | 250,671 | NA | 0.994 (0.967 - 1.022) |
| Ma et al [2024] | A | R7 | Hypertension | 54,358 | 408,652 | NA | 0.991 (0.981 - 1.001) |
| Ma et al [2024] | B | R7 | Hypertension | 54,358 | 408,652 | NA | 0.998 (0.994 - 1.002) |
| Ma et al [2024] | C | R7 | Hypertension | 54,358 | 408,652 | NA | 1.001 (0.999 - 1.002) |
| Zhang et al [2024] | C | R5 | Female infertility | 6,481 | 68,969 | 4 | 0.4701966 (0.1569 - 1.4093) |
| Shen et al [2024] | A | R7 | Liver diseases | 11,934 | 400,247 | 15 | 0.91 (0.82 - 0.99) |
| Shen et al [2024] | A | R7 | Kidney cyst | 1,874 | 408,319 | 18 | 1.14 (0.9 - 1.44) |
| Shen et al [2024] | A | R7 | Other specified disorders of kidney and ureter | 948 | 408,319 | 18 | 1.15 (0.83 - 1.6) |
| Shen et al [2024] | A | R7 | Other disorders of kidney and ureter | 3,862 | 408,319 | 18 | 1.12 (0.97 - 1.31) |
| Shen et al [2024] | A | R7 | Hypothyroidism | 45,321 | 298,847 | 11 | 0.89 (0.8 - 1.01) |
| Shen et al [2024] | A | R7 | Thyroid disorders | 62,464 | 349,717 | 11 | 0.92 (0.85 - 1) |
| Shen et al [2024] | A | R7 | Graves’ disease | 3,176 | 409,005 | 11 | 0.82 (0.66 - 1.02) |
| Zhang et al [2024] | A | R7 | BC | 7,480 | 329,679 | 413 | 0.9917 (0.9865 - 0.997) |
| Zhang et al [2024] | C | R7 | Invasive mucinous ovarian cancer | 1,417 | 40,941 | 3 | 1.0718 (1.021 - 1.1252) |
| Zhang et al [2024] | C | R7 | Rheumatoid arthritis | 1,523 | 461,487 | 11 | 0.9673 (0.9446 - 0.9906) |
| Zhang et al [2024] | B | R7 | MS | 1,679 | 461,254 | 4 | 7.4877E+11 (2153.5032 - 2.60E+20) |
| Zhang et al [2024] | C | R7 | SLE | 538 | 213,145 | 41 | 0.9858 (0.9749 - 0.9967) |
| Zhang et al [2024] | A | R7 | SLE | 538 | 213,145 | 42 | 0.956 (0.9213 - 0.9919) |
| Zhang et al [2024] | B | R7 | SLE | 538 | 213,145 | 41 | 0.9767 (0.9546 - 0.9993) |
| Zhang et al [2024] | C | R7 | Rheumatoid arthritis | 1,523 | 461,487 | 1 | 0.00000188 (6.27E-11 - 0.0561) |
| Zhang et al [2024] | A | R7 | Ulcerative colitis | 4,320 | 210,300 | 7 | 1.2543 (1.0621 - 1.4813) |
| Zhang et al [2024] | C | R7 | T2DM | 77,418 | 356,122 | 2 | 0.000005 (2.36E-10 - 0.1058) |
| Zhang et al [2024] | C | R7 | Diabetic nephropathy | 3,283 | 210,463 | 2 | 0.9492 (0.9145 - 0.9853) |
| Zhang et al [2024] | C | R7 | Glomerulonephritis | 4,613 | 214,179 | 2 | 0.8961 (0.8174 - 0.9824) |
| Fang et al [2024] | C | R5 | Intrahepatic Cholestasis of Pregnancy | 940 | 122,639 | 9 | 1.42 (0.85 - 2.36) |
| Fang et al [2024] | C | R5 | Placental disorders | 102 | 104,247 | 9 | 4.81 (1.05 - 22.05) |
| Fang et al [2024] | C | R5 | Gestational hypertension/pre-eclampsia | 1,864 | 461,069 | 7 | 1.00 (1.00 - 1.00) |
| Fang et al [2024] | C | R5 | Number of spontaneous miscarriages | NA | NA | 8 | 1.00 (0.96 - 1.04) |
| Fang et al [2024] | C | R5 | Number of stillbirths | NA | NA | 9 | 1.00 (0.98 - 1.01) |

A, COVID-19 severity; AAP, Acute alcoholic pancreatitis; ACP, Alcoholic chronic pancreatitis; AD, Alzheimer's disease; ADHD, Attention deficit hyperactivity disorder; AF, Atrial fibrillation; AG, Acute gastritis; ALS, Amyotrophic lateral sclerosis; AMI, Acute myocardial infarction; AP, Acute pancreatitis; AQP4+NMOSD, Aquaporin-4 antibody-positive neuromyelitis optica spectrum disorder; AQP4-NMOSD, Aquaporin-4 antibody-negative neuromyelitis optica spectrum disorder; ASD, Autistic spectrum disorders; ATIN, Acute tubulo-interstitial nephritis; B, COVID-19 hospitalized; BC, Breast cancer; HER, Human epidermal growth factor receptor 2; BD, Bipolar disorder; C, COVID-19 susceptibility; CAD, Coronary artery disease; CD40L, Cluster of differentiation 40 ligand; CES, Cardioembolic stroke; CHD, Coronary heart disease; CKD, Chronic kidney disease; CNS, Central nervous system; CP, Chronic pancreatitis; CRC, Colorectal cancer; CTIN, Chronic tubulo-interstitial nephritis; CVS, Cardiovascular and cerebrovascular diseases; DKD, Diabetic kidney disease; EC, Endometrial cancer; ED, Erectile dysfunction; EOC, Epithelial ovarian cancer; EOMG, Early-onset myasthenia gravis; ER+BC, Estrogen receptor positive breast cancer; ER-BC, Estrogen receptor negative breast cancer; FL, Follicular lymphoma; GAD, Generalized anxiety disorder; GBM, Glioblastoma multiforme; GBS, Guillain-barré syndrome; GRED, Gastroesophageal reflux disease; GS, Gastric ulcer; HBV, Hepatitis b virus; HF, Heart failure; HTN, Hypertension; HL, Hodgkin’s lymphoma; HNC, Head and neck cancer; IBS, Irritable bowel syndrome; IS, Ischemic stroke; LAS, Large artery stroke; LOMG, Late-onset myasthenia gravis; MDD, Major depressive disorder; ME/CFS, Myalgic encephalomyelitis/chronic fatigue syndrome; MG, Myasthenia gravis; mGCIPL, Macular ganglion cell-inner plexiform layer; MI, Myocardial infarction; mRNFL, Macular retinal nerve fiber layer; MS, Multiple sclerosis; NA, Not applicable; NAFLD, Nonalcoholic fatty liver disease; NHL DLBCL, Non-hodgkin lymphoma, diffuse large b-cell lymphoma; NMOSD, Neuromyelitis optica spectrum disorders; NRVHD, Non-rheumatic valvular heart disease; OCD, Obsessive compulsive disorder; PAD, Peripheral artery disease; PCOS, Polycystic ovary syndrome; PD, Parkinson's disease; PE, Pulmonary embolism; PTSD, Post-traumatic stress disorder; RA, Rheumatoid arthritis; RHD, Rheumatic heart disease; SCZ, Schizophrenia; SLE, Systemic lupus erythematosus; SVS, Small vessel stroke; T1DM, Type 1 diabetes mellitus; T2DM, Type 2 diabetes mellitus; TIA, Transient ischemic attack; TIN, Tubulointerstitial nephritis; TNFR1, Tumor necrosis factor receptor 1; TNFSF14, Tumor necrosis factor ligand superfamily member 14; TRAIL, Tumor necrosis factor-related apoptosis-inducing ligand; TS, Tourette’s syndrome; Tu, Tumour; UC, Ulcerative colitis; VTE, Venous thrombus embolism.

eTable 2 Quality assessment of the included studies.

| Author, year | Title and Abstract | Objectives | Study Design and Data Sources | MR Assumption | | | Analytical Methods | Sensitivity and Additional Analyses | Participant Summary Statistics | Main MR Results | Addresses Limitations | Interprets MR Results | Power Calculations | Discusses Generaliz-ability of Results |
| --- | --- | --- | --- | --- | --- | --- | --- | --- | --- | --- | --- | --- | --- | --- |
|  |  |  |  | 1 | 2 | 3 |  |  |  |  |  |  |  |  |
| Sun, et al. ,2023 | Yes | Yes | Yes | Yes | Yes | Yes | Yes | Yes | Yes | Yes | Partial Yes | Yes | Partial Yes | No |
| Ding et al. ,2023 | Yes | Yes | Yes | Partial Yes | Yes | Yes | Yes | Yes | Yes | Yes | Yes | Yes | Partial Yes | Yes |
| Zhang et al. ,2022 | Yes | Yes | Yes | Yes | Yes | Yes | Yes | Yes | Yes | Yes | Yes | Yes | Yes | No |
| Tirozzi et al. ,2022 | Yes | Yes | Partial Yes | Yes | Yes | Yes | Yes | Yes | Yes | Yes | Yes | Yes | Partial Yes | Yes |
| Dai et al. ,2024 | Yes | Yes | Yes | Yes | Yes | Yes | Yes | Yes | Yes | Yes | Yes | Yes | Partial Yes | Yes |
| Chen et al. ,2023 | Yes | Yes | Yes | Partial Yes | Yes | Yes | Yes | Yes | Yes | Yes | Yes | Yes | Partial Yes | Yes |
| Baranova et al. ,2022 | Yes | Yes | Yes | Yes | Yes | Yes | Yes | Yes | Yes | Yes | Yes | Yes | Yes | Partial Yes |
| Ran et al. ,2023 | Yes | Yes | Yes | Yes | Yes | Yes | Yes | Yes | Yes | Yes | No | Yes | Partial Yes | No |
| Cao et al. ,2023 | Yes | Yes | Partial Yes | Yes | Yes | Yes | Yes | Yes | Yes | Yes | Yes | Yes | Partial Yes | Yes |
| Zhu et al. ,2022 | Yes | Yes | Yes | Yes | Yes | Yes | Yes | Yes | Yes | Yes | Yes | Yes | Partial Yes | Yes |
| Li et al. ,2023 | Yes | Yes | Yes | Yes | Yes | Yes | Yes | Yes | Yes | Yes | Partial Yes | Yes | Partial Yes | No |
| Baranova et al. ,2023 | Yes | Yes | Yes | Yes | Yes | Yes | Yes | Yes | Yes | Yes | Yes | Yes | Partial Yes | Yes |
| Xue et al. ,2023 | Yes | Yes | Yes | Yes | Yes | Yes | Yes | Yes | Yes | Yes | Partial Yes | Yes | Partial Yes | Yes |
| Pan et al. ,2024 | Yes | Yes | Yes | Yes | Yes | Yes | Yes | Yes | Yes | Yes | Yes | Yes | Partial Yes | Yes |
| Xing et al. ,2023 | Yes | Yes | Yes | Yes | Yes | Yes | Yes | Yes | Yes | Yes | Yes | Yes | Partial Yes | Yes |
| You et al. ,2023 | Yes | Yes | Yes | Yes | Yes | Yes | Yes | Yes | Yes | Yes | Yes | Yes | Partial Yes | Yes |
| He et al. ,2023 | Yes | Yes | Yes | Yes | Yes | Yes | Yes | Yes | Yes | Yes | Yes | Yes | Yes | Yes |
| Tan et al. ,2021 | Yes | Yes | Yes | Yes | Yes | Yes | Yes | Yes | Yes | Yes | Yes | Yes | Partial Yes | Yes |
| Zhang et al. ,2022 | Yes | Yes | Yes | Yes | Yes | Yes | Yes | Yes | Yes | Yes | Yes | Yes | Yes | Yes |
| Tan et al. ,2022 | Yes | Yes | Yes | Yes | Yes | Yes | Yes | Yes | Yes | Yes | Yes | Yes | Partial Yes | Yes |
| Zuber et al. ,2021 | Yes | Yes | Yes | Partial Yes | Yes | Yes | Yes | Yes | Yes | Yes | Partial Yes | Yes | Partial Yes | No |
| Zhang et al. ,2022 | Yes | Yes | Yes | Yes | Yes | Yes | Yes | Yes | Yes | Yes | Yes | Yes | Yes | Partial Yes |
| Huang et al. ,2023 | Yes | Yes | Yes | Yes | Yes | Yes | Yes | Yes | Yes | Yes | Yes | Yes | Yes | Partial Yes |
| Wang et al. ,2022 | Yes | Yes | Yes | Yes | Yes | Yes | Yes | Yes | Yes | Yes | Partial Yes | Yes | Partial Yes | Yes |
| Zheng et al. ,2023 | Yes | Yes | Yes | Yes | Yes | Yes | Yes | Yes | Yes | Yes | Yes | Yes | Partial Yes | Yes |
| Wang et al. ,2023 | Yes | Yes | Yes | Yes | Yes | Yes | Yes | Yes | Yes | Yes | Partial Yes | Yes | Partial Yes | Yes |
| Jia et al. ,2022 | Yes | Yes | Yes | Yes | Yes | Yes | Yes | Yes | Yes | Yes | Yes | Yes | Partial Yes | Yes |
| Liu et al. ,2023 | Yes | Yes | Yes | Partial Yes | Yes | Yes | Yes | Yes | Yes | Yes | Yes | Yes | Partial Yes | Yes |
| Ma et al. ,2023 | Yes | Yes | Yes | Yes | Yes | Yes | Yes | Yes | Yes | Yes | Partial Yes | Yes | Partial Yes | Yes |
| Li et al. ,2024 | Yes | Yes | Yes | Yes | Yes | Yes | Yes | Yes | Yes | Yes | Partial Yes | Yes | Partial Yes | Yes |
| Ong et al. ,2021 | Partial Yes | Yes | Yes | Yes | Yes | Yes | Yes | Yes | Yes | Yes | Partial Yes | Yes | No | Yes |
| Wang et al. ,2023 | Yes | Yes | Yes | Yes | Yes | Yes | Yes | Yes | Yes | Yes | Partial Yes | Yes | Partial Yes | Yes |
| Ge et al. ,2023 | Yes | Yes | Yes | Partial Yes | Yes | Yes | Yes | Yes | Yes | Yes | No | No | Yes | Yes |
| Liu et al. ,2021 | Partial Yes | Yes | Yes | Yes | Yes | Yes | Yes | Yes | Yes | Yes | Yes | Yes | Yes | Yes |
| Cao et al. ,2022 | Yes | Yes | Yes | Yes | Yes | Yes | Yes | Yes | Yes | Yes | Yes | Partial Yes | Partial Yes | Yes |
| Li et al. ,2022 | Yes | Yes | Yes | Yes | Yes | Yes | Yes | Yes | Yes | Yes | No | Yes | Partial Yes | Yes |
| Yao et al. ,2023 | Yes | Yes | Yes | Yes | Yes | Yes | Yes | Yes | Yes | Yes | Yes | Yes | Partial Yes | Yes |
| Ran et al. ,2022 | Yes | Yes | Yes | Partial Yes | Yes | Yes | Yes | Yes | Yes | Yes | No | No | Partial Yes | No |
| Yang et al. ,2023 | Yes | Yes | Yes | Yes | Yes | Yes | Yes | Yes | Yes | Yes | Yes | Yes | Partial Yes | No |
| Xu et al. ,2023 | Yes | Yes | Yes | Yes | Yes | Yes | Yes | Yes | Yes | Yes | No | Yes | Partial Yes | Yes |
| Sun et al. ,2023 | Yes | Yes | Yes | Yes | Yes | Yes | Yes | Yes | Yes | Yes | No | Yes | Partial Yes | Yes |
| Luo et al. ,2023 | Yes | Yes | Yes | Yes | Yes | Yes | Yes | Yes | Yes | Yes | No | Yes | Partial Yes | Yes |
| Chalitsios et al. ,2022 | Yes | Yes | Yes | Yes | Yes | Yes | Yes | Yes | Yes | Yes | No | Yes | Partial Yes | Yes |
| Quan et al. ,2023 | Yes | Yes | Yes | Yes | Yes | Yes | Yes | Yes | Yes | Yes | No | Yes | Partial Yes | Yes |
| Sun et al. ,2023 | Yes | Yes | Yes | Yes | Yes | Yes | Yes | Yes | Yes | Yes | Partial Yes | Yes | Yes | Yes |
| Wang et al. ,2023 | Yes | Yes | Yes | Yes | Yes | Yes | Yes | Yes | Yes | Yes | Yes | Yes | Partial Yes | Yes |
| Peng et al. ,2022 | Yes | Yes | Yes | Yes | Yes | Yes | Yes | Yes | Yes | Yes | No | Yes | Partial Yes | No |
| Wang et al. ,2023 | Yes | Yes | Yes | Yes | Yes | Yes | Yes | Yes | Yes | Yes | No | Yes | Partial Yes | Yes |
| Chen et al. ,2023 | Yes | Yes | Yes | Yes | Yes | Yes | Yes | Yes | Yes | Yes | No | Yes | Partial Yes | Yes |
| Dong et al. ,2023 | Yes | Yes | Yes | Yes | Yes | Yes | Yes | Yes | Yes | Yes | No | Yes | Partial Yes | Yes |
| Song et al. ,2023 | Yes | Yes | Yes | Yes | Yes | Yes | Yes | Yes | Yes | Yes | No | Yes | Partial Yes | Yes |
| Li et al. ,2023 | Yes | Yes | Yes | Yes | Yes | Yes | Yes | Yes | Yes | Yes | No | Yes | Partial Yes | Yes |
| Zhao et al. ,2023 | Yes | Yes | Yes | Yes | Yes | Yes | Yes | Yes | Yes | Yes | Yes | Yes | Yes | No |
| Sun et al. ,2023 | Yes | Yes | Yes | Yes | Yes | Yes | Yes | Yes | Yes | Yes | No | Yes | Partial Yes | No |
| Xu et al. ,2023 | Yes | Yes | Yes | Yes | Yes | Yes | Yes | Yes | Yes | Yes | No | Yes | Partial Yes | No |
| Wu et al. ,2022 | Yes | Yes | Yes | Yes | Yes | Yes | Yes | Yes | Yes | Yes | Yes | Yes | Partial Yes | Yes |
| Lu et al. ,2023 | Yes | Yes | Yes | Yes | Yes | Yes | Yes | Yes | Yes | Yes | Yes | Yes | Partial Yes | Yes |
| Liu et al. ,2023 | Yes | Yes | Yes | Yes | Yes | Yes | Yes | Yes | Yes | Yes | No | Yes | Partial Yes | Yes |
| Song et al. ,2023 | Yes | Yes | Yes | Yes | Yes | Yes | Yes | Yes | Yes | Yes | Yes | Yes | Partial Yes | Yes |
| Shi et al. ,2022 | Yes | Yes | Yes | Yes | Yes | Yes | Yes | Yes | Yes | Yes | No | Yes | Partial Yes | Yes |
| Chang et al. ,2023 | Yes | Yes | Yes | Yes | Yes | Yes | Yes | Yes | Yes | Yes | Yes | Yes | Partial Yes | No |
| Zhang et al. ,2022 | Yes | Yes | Yes | Yes | Yes | Yes | Yes | Yes | Yes | Yes | No | Yes | Partial Yes | Yes |
| Si et al. ,2023 | Yes | Yes | Yes | Yes | Yes | Yes | Yes | Yes | Yes | Yes | Yes | Yes | Partial Yes | Yes |
| Liu et al. ,2024 | Yes | Yes | Yes | Yes | Yes | Yes | Yes | Yes | Yes | Yes | No | Yes | Partial Yes | No |
| Lin et al. ,2024 | Yes | Yes | Yes | Yes | Yes | Yes | Yes | Yes | Yes | Yes | Yes | Yes | Partial Yes | Yes |
| Tang et al. ,2024 | Yes | Yes | Yes | Yes | Yes | Yes | Yes | Yes | Yes | Yes | Yes | Yes | Yes | Yes |
| Wu et al. ,2024 | Yes | Yes | Yes | Yes | Yes | Yes | Yes | Yes | Yes | Yes | Yes | Yes | Yes | Yes |
| Baranova et al. ,2024 | Yes | Yes | Yes | Yes | Yes | Yes | Yes | Yes | Yes | Yes | Yes | Yes | Yes | Yes |
| Cao et al. ,2024 | Yes | Yes | Yes | Yes | Yes | Yes | Yes | Yes | Yes | Yes | Yes | Yes | Yes | Yes |
| Jiang et al. ,2024 | Yes | Yes | Yes | Yes | Yes | Yes | Yes | Yes | Yes | Yes | Yes | Yes | Yes | Yes |
| Monistrol-Mula et al. ,2024 | Yes | Yes | Yes | Yes | Yes | Yes | Yes | Yes | Yes | Yes | Yes | Yes | Partial Yes | Yes |
| Wang et al. ,2024 | Yes | Yes | Yes | Yes | Yes | Yes | Yes | Yes | Yes | Yes | Yes | Yes | Yes | Yes |
| Tang et al. ,2024 | Yes | Yes | Yes | Yes | Yes | Yes | Yes | Yes | Yes | Yes | Yes | Yes | Yes | Yes |
| Li et al. ,2024 | Yes | Yes | Yes | Yes | Yes | Yes | Yes | Yes | Yes | Yes | Yes | Yes | Yes | Yes |
| He et al. ,2024 | Yes | Yes | Yes | Yes | Yes | Yes | Yes | Yes | Yes | Yes | Yes | Yes | Yes | Yes |
| Zou et al. ,2024 | Yes | Yes | Yes | Yes | Yes | Yes | Yes | Yes | Yes | Yes | Yes | Yes | Yes | Yes |
| Kong et al. ,2024 | Yes | Yes | Yes | Yes | Yes | Yes | Yes | Yes | Yes | Yes | Yes | Yes | Yes | Yes |
| Wang et al. ,2024 | Yes | Yes | Yes | Yes | Yes | Yes | Yes | Yes | Yes | Yes | Yes | Yes | Yes | Yes |
| Gao et al. ,2024 | Yes | Yes | Yes | Yes | Yes | Yes | Yes | Yes | Yes | Yes | Yes | Yes | Yes | Yes |
| Wang et al. ,2024 | Yes | Yes | Yes | Yes | Yes | Yes | Yes | Yes | Yes | Yes | Yes | Yes | Yes | Yes |
| Han et al. ,2024 | Yes | Yes | Yes | Yes | Yes | Yes | Yes | Yes | Yes | Yes | Yes | Yes | Yes | Yes |
| Song et al. ,2024 | Yes | Yes | Yes | Yes | Yes | Yes | Yes | Yes | Yes | Yes | Yes | Yes | Yes | Yes |
| Ma et al. ,2024 | Yes | Yes | Yes | Yes | Yes | Yes | Yes | Yes | Yes | Yes | Yes | Yes | Yes | Yes |
| Zhang et al. ,2024 | Yes | Yes | Yes | Yes | Yes | Yes | Yes | Yes | Yes | Yes | Yes | Yes | Yes | Yes |
| Shen et al. ,2024 | Yes | Yes | Yes | Yes | Yes | Yes | Yes | Yes | Yes | Yes | Yes | Yes | Yes | Yes |
| Zhang et al. ,2024 | Yes | Yes | Yes | Yes | Yes | Yes | Yes | Yes | Yes | Yes | Yes | Yes | Yes | Yes |
| Fang et al. ,2024 | Yes | Yes | Yes | Yes | Yes | Yes | Yes | Yes | Yes | Yes | Yes | Yes | Partial Yes | Yes |

For quality assessment, information was extracted based on a template developed from the Strengthening the Reporting of Observational Studies in Epidemiology using Mendelian Randomization (STROBE-MR) guidelines.In the quality assessment table, yes represents that the study has fully addressed the specific aspect, with high quality and rigorous methodology, ensuring reliable results. Partial Yes indicates that the study has partially addressed the aspect, but lacks sufficient detail or clarity, suggesting some uncertainty or potential limitations. No that the study has either not addressed the aspect at all or has done so poorly, resulting in a significant risk of bias and lower overall reliability.

eTable 3 Mendelian Randomization meta-analysis of research findings

| **CNS** | | | | | | | **CVS** | | | | | | |
| --- | --- | --- | --- | --- | --- | --- | --- | --- | --- | --- | --- | --- | --- |
| **Disease** | **First author [year]** | **SS** | **I^2^** | **OR** | **Lower** | **Upper** | **Disease** | **First author [year]** | **SS** | **I^2^** | **OR** | **Lower** | **Upper** |
| AD | Dai et al [2024] Sun et al [2023] Ding et al [2023] | 10 | 27.10% | 1.005 | 0.999 | 1.011 | Arrhythmology | Jia et al [2022] Tan et al [2022] Zhang et al [2022] Li et al [2024] | 10 | 36.50% | 1.009 | 0.986 | 1.032 |
| ADHD | Chen et al [2023] | 3 | 0.00% | 0.999 | 0.991 | 1.007 | CAD | Tan et al [2022] Jia et al [2022] | 5 | 0.00% | 1.009 | 0.992 | 1.026 |
| ALS | Zhang et al [2022] | 3 | 76.00% | 0.934 | 0.774 | 1.094 | Cardiac arrest | Tang et al [2024] | 6 | 0.00% | 0.963 | 0.894 | 1.031 |
| ASD | Chen et al [2023] Xue et al [2023] | 6 | 0.00% | 0.983 | 0.961 | 1.005 | Cerebral infarction | Dai et al [2024] | 3 | 9.60% | 1.002 | 0.994 | 1.011 |
| BD | Xue et al [2023] Monistrol-Mula et al [2024] | 5 | 63.40% | 1.072 | 0.973 | 1.170 | CHD | Zheng et al [2023] | 1 | NA | **1.010** | **1.005** | **1.015** |
| Cognitive  dysfunction | Ran et al [2023] | 3 | 65.30% | **0.860** | **0.774** | **0.945** | HF | Jia et al [2022] Wang et al [2022] | 4 | 12.20% | 1.020 | 0.991 | 1.050 |
| Epilepsy | Dai et al [2024] He et al [2023] Wu et al [2024] Xing et al [2023] You et al [2023] | 32 | 68.30% | **1.087** | **1.053** | **1.122** | HTN | Ma et al [2024] Jia et al [2022] Tan [2021] | 4 | 78.20% | 1.004 | 0.987 | 1.021 |
| GAD | Tirozzi et al [2022] Xue et al [2023] | 6 | 0.00% | **1.010** | **1.006** | **1.015** | Intracerebral hemorrhage | Dai et al [2024] | 3 | 0.00% | 0.987 | 0.958 | 1.016 |
| Insomnia | Ran et al [2023] | 3 | 0.00% | 1.000 | 0.996 | 1.005 | MI | Zheng et al [2023]  Li et al [2024] | 7 | 9.40% | **0.978** | **0.962** | **0.993** |
| Intelligence | Cao et al [2023] Zhu et al [2022] | 3 | 79.80% | 0.963 | 0.925 | 1.002 | Myocarditis | Li et al [2024] Liu et al [2023] | 3 | 0.00% | 0.997 | 0.905 | 1.089 |
| MDD | Baranova et al [2023] Dai et al [2024] Tirozzi et al [2022] Xue et al [2023] Li et al [2023] Monistrol-Mula et al [2024] | 17 | 0.00% | 1.000 | 0.996 | 1.003 | PAD | Jia et al [2022] | 1 | NA | 1.059 | 0.905 | 1.238 |
| MS | Zhang et al [2022] | 3 | 84.50% | 0.993 | 0.856 | 1.131 | PE | Jia et al [2022] | 1 | NA | 0.989 | 0.898 | 1.089 |
| Neurosurgical diseases | Dai et al [2024] Pan et al [2024] | 33 | 0.00% | 1.008 | 0.998 | 1.018 | Pericarditis | Liu et al [2023] | 3 | 0.00% | **0.900** | **0.825** | **0.975** |
| OCD | Dai et al [2024] | 3 | 0.00% | 1.017 | 0.975 | 1.060 | Stroke | Dai et al [2024] Zhang et al [2022] Ma et al [2023] Zuber et al [2021] | 16 | 39.20% | **1.031** | **1.018** | **1.043** |
| Optic nerve | Pan et al [2024] Cao et al [2024] | 5 | 51.70% | **1.444** | **1.080** | **1.807** | Subarachnoid hemorrhage | Dai et al [2024] | 3 | 0.00% | 0.999 | 967.000 | 1.030 |
| PD | Dai et al [2024] Tirozzi et al [2022] | 6 | 0.00% | 0.994 | 0.975 | 1.012 | VHD | Jia et al [2022] | 2 | 0.00% | 1.011 | 0.938 | 1.083 |
| PTSD | Baranova et al [2023] Monistrol-Mula et al [2024] | 4 | 0.00% | 1.000 | 0.995 | 1.004 | VTE | Li et al [2024] Huang et al [2023] Jia et al [2022] | 7 | 79.60% | 1.028 | 0.996 | 1.060 |
| SCZ | Tirozzi et al [2022] Xue et al [2023] Baranova et al [2022] | 8 | 37.60% | **1.048** | **1.014** | **1.081** |  |  |  |  |  |  |  |
| TS | Chen et al [2023] | 3 | 0.00% | 0.998 | 0.978 | 1.017 |  |  |  |  |  |  |  |
| **DS** | | | | | | | **GU** | | | | | | |
| **Disease** | **First author [year]** | **SS** | **I^2^** | **OR** | **Lower** | **Upper** | **Disease** | **First author**  **[year]** | **SS** | **I^2^** | **OR** | **Lower** | **Upper** |
| AAP | Ge et al [2023] | 6 | 45.20% | 0.996 | 0.949 | 1.044 | CKD | Chang et al [2023] Lin et al [2024] | 4 | 0.00% | 1.005 | 0.961 | 1.049 |
| ACP | Ge et al [2023] | 6 | 18.40% | 0.973 | 0.945 | 1.001 | DKD | Chang et al [2023] | 6 | 0.00% | 0.962 | 0.876 | 1.047 |
| Acute  Appendicitis | Wang et al [2023] | 3 | 0.80% | 0.947 | 0.864 | 1.031 | ED | Chang et al [2023] Ma et al [2024] Zhang et al [2022] | 7 | 0.00% | 1.035 | 0.996 | 1.074 |
| AG | Wang et al [2023] | 2 | 0.00% | 1.000 | 0.995 | 1.005 | Female infertility | Zhang et al [2024] | 1 | NA | 0.470 | 0.160 | 1.410 |
| AP | Ge et al [2023] Wang et al [2023] | 9 | 9.60% | 0.996 | 0.954 | 1.039 | Genital infection | Chang et al [2023] | 3 | 0.00% | 0.983 | 0.907 | 1.059 |
| Barrett's  esophagus | He et al [2024] | 2 | 0.00% | 1.084 | 0.963 | 1.205 | Hydrocele | Chang et al [2023] | 3 | 0.00% | 1.022 | 0.940 | 1.103 |
| Chlocystitis | Wang et al [2023] | 3 | 41.00% | 1.048 | 0.889 | 1.208 | Hydronephrosis | Chang et al [2023] | 3 | 0.00% | 0.957 | 0.829 | 1.085 |
| Cholangitis | Wang et al [2023] | 3 | 21.90% | 0.969 | 0.823 | 1.116 | Hypertensive Renal Disease | Chang et al [2023] | 3 | 0.00% | 0.940 | 0.758 | 1.122 |
| Cholelithiasis | Wang et al [2023] | 3 | 0.00% | 0.993 | 0.848 | 1.138 | Male infertility | Liu et al [2024] | 2 | 0.00% | 0.907 | 0.764 | 1.050 |
| Chronic  Gastritis | Wang et al [2023] | 3 | 0.00% | 0.958 | 0.787 | 1.130 | Miscarriage | Shi et al [2022] | 2 | 0.00% | 1.000 | 0.996 | 1.004 |
| Congenital  esophagea | He et al [2024] | 2 | 0.00% | 1.514 | 0.977 | 2.051 | Number of stillbirths | Fang et al [2024] | 1 | NA | 1.000 | 0.985 | 1.015 |
| CP | Ge et al [2023] Wang et al [2023] | 9 | 0.00% | 0.998 | 0.978 | 1.019 | Orchitis and epididymitis | Chang et al [2023] | 3 | 0.00% | 1.029 | 0.901 | 1.156 |
| CRC | Wang et al [2023] | 3 | 0.00% | 1.004 | 0.976 | 1.032 | Other disorders of kidney and ureter | Shen et al [2024] | 1 | NA | 1.120 | 0.950 | 1.290 |
| Crohn | Wang et al [2023] | 3 | 0.00% | 1.042 | 0.983 | 1.101 | Other specified disorders of kidney and ureter | Shen et al [2024] | 1 | NA | 1.150 | 0.765 | 1.535 |
| Esophageal  ulcer | He et al [2024] | 2 | 0.00% | 1.010 | 0.924 | 1.096 | PCOS | Si et al [2023] | 3 | 0.00% | 0.963 | 0.874 | 1.051 |
| Esophageal  varices | He et al [2024] | 2 | 0.00% | 1.081 | 0.960 | 1.201 | Placental disorders | Fang et al [2024] | 1 | NA | **4.810** | **1.050** | **22.050** |
| Esophagitis | He et al [2024] | 2 | 0.00% | 1.051 | 0.947 | 1.155 | Prostatitis | Chang et al [2023] | 3 | 0.00% | 0.947 | 0.852 | 1.042 |
| Gastroesophageal  ref | He et al [2024] | 2 | 0.00% | 0.993 | 0.968 | 1.018 | Renal failure | Chang et al [2023] | 2 | 0.00% | 1.001 | 0.941 | 1.062 |
| GRED | Ong et al [2021] Wang et al [2023] | 4 | 64.90% | 1.010 | 0.982 | 1.039 | Sexual dysfunction | Chang et al [2023] | 3 | 0.00% | **1.093** | **1.026** | **1.160** |
| GS | Wang et al [2023] | 1 | NA | 1.020 | 1.000 | 1.040 | Testicular dysfunction | Chang et al [2023] | 3 | 0.00% | **1.326** | **1.095** | **1.557** |
| IBS | Wang et al [2023] | 3 | 0.00% | **1.039** | **1.014** | **1.063** | TIN | Chang et al [2023] | 8 | 0.00% | 0.977 | 0.947 | 1.007 |
| Intrahepatic  Cholest | Fang et al [2024] | 1 | NA | 1.420 | 0.665 | 2.175 | Torsion of testis | Chang et al [2023] | 3 | 0.00% | **0.772** | **0.554** | **0.990** |
| Liver  diseases | Shen et al [2024] | 1 | NA | 0.910 | 0.825 | 0.995 | Urolithiasis | Chang et al [2023] | 3 | 0.00% | **1.293** | **1.043** | **1.543** |
| NAFLD | Liu et al [2021] Wang et al [2023] | 6 | 0.00% | 0.956 | 0.899 | 1.013 |  |  |  |  |  |  |  |
| Oesophageal obstruct | He et al [2024] | 2 | 0.00% | 0.914 | 0.809 | 1.019 |  |  |  |  |  |  |  |
| Perforation of esoph | He et al [2024] | 2 | 0.00% | 1.113 | 0.724 | 1.502 |  |  |  |  |  |  |  |
| UC | Wang et al [2023] | 3 | 58.50% | 1.037 | 0,906 | 1.169 |  |  |  |  |  |  |  |
| **Tu** | | | | | | | **Autoimmune disease** | | | | | | |
| **Disease** | **First author [year]** | **SS** | **I^2^** | **OR** | **Lower** | **Upper** | **Disease** | **First author [year]** | **SS** | **I^2^** | **OR** | **Lower** | **Upper** |
| BC | Zhao et al [2023] Li et al [2023] | 14 | 6.10% | 1.006 | 0.992 | 1.019 | Ankylosing spondylitis | Quan et al [2023] | 1 | NA | 1.073 | 0.833 | 1.381 |
| Benign esophageal tumors | He et al [2024] | 2 | 0.00% | 0.854 | 0.673 | 1.036 | Atopic dermatitis | Luo et al [2023] | 2 | 0.00% | 1.037 | 0.955 | 1.118 |
| Benign meningioma | Dai et al [2024] | 3 | 0.00% | 0.989 | 0.944 | 1.034 | Celiac disease | Zou et al [2024] | 3 | 0.00% | **1.111** | **1.011** | **1.211** |
| Benign neoplasm of brain and other parts of CNS | Dai et al [2024] | 3 | 0.00% | 0.997 | 0.947 | 1.048 | GBS | Kong et al [2024] | 1 | NA | 0.830 | 0.600 | 1.160 |
| Benign neoplasm:  Adrenal gland | Chang et al [2023] | 3 | 0.00% | 1.071 | 0.921 | 1.231 | Gout | Quan et al [2023] | 1 | NA | 1.075 | 0.960 | 1.204 |
| Benign neoplasm: Bladder | Chang et al [2023] | 3 | 0.00% | 0.903 | 0.533 | 1.273 | Juvenile idiopathic arthritis | Quan et al [2023] | 1 | NA | **1.517** | **1.144** | **2.011** |
| Benign neoplasm: Kidney | Chang et al [2023] | 3 | 0.00% | 0.921 | 0.730 | 1.112 | MG | Sun et al [2023] | 9 | 0.00% | **1.096** | **1.029** | **1.162** |
| Benign neoplasm: prostate | Chang et al [2023] | 3 | 0.00% | 0.742 | 0.427 | 1.057 | NMOSD | Sun et al [2023] Wang et al [2023] | 12 | 0.00% | **1.292** | **1.099** | **1.484** |
| Brain  glioblastoma cancer | Li et al [2023]) | 3 | 0.00% | 0.924 | 0.566 | 1.282 | Primary biliary cholangitis | Quan et al [2023] | 1 | NA | **1.370** | **1.150** | **1.640** |
| Cerebral aneurysm | Dai et al [2024] | 3 | 0.00% | 0.990 | 0.960 | 1.020 | Primarys jogren's syndrome | Quan et al [2023] | 1 | NA | 0.995 | 0.796 | 1.244 |
| Colon cancer | Jin et al [2023] | 3 | 0.00% | 1.061 | 0.964 | 1.158 | Psoriasis | Chalitsios et al [2022] | 3 | 0.00% | 0.961 | 0.796 | 1.126 |
| Colorectal cancer | Jin et al [2023] Li et al [2023] | 4 | 0.00% | 1.000 | 0.995 | 1.005 | RA | Yao et al [2023]  Zhang et al [2023] | 4 | 0.00% | **0.976** | **0.954** | **0.999** |
| Corpus uteri cancer | Li et al [2023]) | 3 | 0.00% | 0.891 | 0.777 | 1.006 | SLE | Ran et al [2022] Quan et al [2023] Yang et al [2023] Yao et al [2023] Zhang et al [2024] | 16 | 20.40% | 0.982 | 0.962 | 1.002 |
| EC | Zhao et al [2023] Wu et al [2022] | 6 | 0.00% | **1.083** | **1.047** | **1.120** | T1DM | Wang et al [2024] | 1 | NA | 1.160 | 0.850 | 1.590 |
| EOC | Zhao et al [2023] | 3 | 0.00% | 1.047 | 0.993 | 1.101 | Zoster | Gao et al [2024] | 3 | 0.00% | 0.965 | 0.863 | 1.067 |
| Esophageal adenocarcinoma | He et al [2024] | 2 | 0.00% | 1.010 | 0.925 | 1.095 | **IDs** | | | | | | |
| Esophageal cancer | Jin et al [2023] | 3 | 0.00% | 0.974 | 0.724 | 1.223 | **Disease** | **First author [year]** | **SS** | **I^2^** | **OR** | **Lower** | **Upper** |
| Eye and  adnexa cancer | Li et al [2023] | 3 | 0.00% | 1.114 | 0.809 | 1.419 | HBV | Liu et al [2023] | 3 | 0.00% | 1.002 | 0.948 | 1.055 |
| Gastric carcinoma | Wang et al [2023] | 2 | 29.70% | 0.980 | 0.925 | 1.036 | Periodontitis | Song et al [2023] | 3 | 0.00% | 1.011 | 0.934 | 1.088 |
| GBM | Dong et al [2023] Dai et al [2024] | 6 | 0.00% | **1.085** | **1.001** | **1.170** | Sepsis | Lu et al [2023] | 3 | 0.00% | 0.988 | 0.953 | 1.023 |
| Genitourinary benign neoplasm | Chang et al [2023] | 1 | NA | 1.140 | 0.640 | 1.640 | **ES** | | | | | | |
| Haemotological cancer | Li et al [2023] | 1 | NA | 0.990 | 0.805 | 1.175 | **Disease** | **First author [year]** | **SS** | **I^2^** | **OR** | **Lower** | **Upper** |
| Heart, mediastinum and pleura cancer | Li et al [2023] | 3 | 0.00% | 0.860 | 0.563 | 1.158 | Autoimmune thyroiddisease | Li et al [2022] | 3 | 0.00% | 1.004 | 0.992 | 1.017 |
| Kidney, except renal pelvis cancer | Li et al [2023] | 3 | 0.00% | 0.952 | 0.845 | 1.059 | Hyperthyroidism | Li et al [2022] | 3 | 0.00% | 0.990 | 0.952 | 1.029 |
| Laryngeal cancer | Wang et al [2024]  Li et al [2023] | 6 | 0.00% | **0.553** | **0.368** | **0.737** | Hypothyroidism | Li et al [2022] | 4 | 83.70% | 1.051 | 0.953 | 1.150 |
| Lip, oral cavity and pharynx cancer | Li et al [2023] | 3 | 0.00% | **0.740** | **0.507** | **0.973** | T2DM | Cao et al [2022] Zhang et al [2024] | 7 | 49.50% | **1.024** | **0.987** | **1.062** |
| Lung adenocarcinoma | Song et al [2024] | 1 | NA | 0.910 | 0.620 | 1.330 | Graves' disease | Shen et al [2024] | 1 | NA | 0.820 | 0.660 | 1.020 |
| Lung cancer | Song et al [2024] | 1 | NA | 1.010 | 0.780 | 1.300 | Thyroid disorders | Shen et al [2024] | 1 | NA | 0.920 | 0.850 | 1.000 |
| Lung squamous cell carcinoma | Song et al [2024] | 1 | NA | 0.840 | 0.570 | 1.240 |  |  |  |  |  |  |  |
| Lymphoma | Chen et al [2023] | 5 | 0.00% | 1.208 | 0.997 | 1.420 |  |  |  |  |  |  |  |
| Malignant meningioma | Dai et al [2024] | 3 | 0.00% | 0.989 | 0.929 | 1.049 |  |  |  |  |  |  |  |
| Malignant neoplasm of brain and other parts of CNS | Dai et al [2024] | 3 | 0.00% | 1.055 | 0.932 | 1.178 |  |  |  |  |  |  |  |
| Malignant neoplasm of testis | Chang et al [2023] | 2 | 0.00% | 0.829 | 0.580 | 1.077 |  |  |  |  |  |  |  |
| Meninges cancer | Li et al [2023] | 3 | 0.00% | 1.021 | 0.869 | 1.173 |  |  |  |  |  |  |  |
| Non-small  cell lung cancer | Li et al [2023] | 3 | 0.00% | 1.011 | 0.922 | 1.100 |  |  |  |  |  |  |  |
| Oesophageal cancer | Li et al [2023] | 1 | NA | 1.030 | 0.644 | 1.648 |  |  |  |  |  |  |  |
| Ovarian cancer | Li et al [2023] | 1 | NA | 1.002 | 0.999 | 1.005 |  |  |  |  |  |  |  |
| Pancreas cancer | Li et al [2023] | 3 | 0.00% | 1.058 | 0.913 | 1.203 |  |  |  |  |  |  |  |
| Prostate cancer | Li et al [2023] | 3 | 0.00% | 1.003 | 0.971 | 1.035 |  |  |  |  |  |  |  |
| Small cell  lung cancer | Li et al [2023] Song et al [2024] | 4 | 9.8%% | 0.836 | 0.619 | 1.052 |  |  |  |  |  |  |  |
| Stomach cancer | Jin et al [2023] Li et al [2023] | 6 | 0.00% | **1.270** | **1.129** | **1.410** |  |  |  |  |  |  |  |
| Testis cancer | Li et al [2023] | 3 | 0.00% | 0.795 | 0.591 | 0.999 |  |  |  |  |  |  |  |
| Thyroid cancer | Li et al [2023] Xu et al [2023] | 6 | 39.90% | 0.913 | 0.745 | 1.081 |  |  |  |  |  |  |  |
| Urological benign neoplasm | Chang et al [2023] | 2 | 0.00% | 0.977 | 0.867 | 1.087 |  |  |  |  |  |  |  |
| Urological malignant neoplasm | Chang et al [2023] | 1 | NA | 1.000 | 0.960 | 1.040 |  |  |  |  |  |  |  |
| Vulva cancer | Li et al [2023] | 3 | 0.00% | 0.933 | 0.634 | 1.231 |  |  |  |  |  |  |  |

AAP, Acute alcoholic pancreatitis; ACP, Alcoholic chronic pancreatitis; AD, Alzheimer's disease; ADHD, Attention deficit hyperactivity disorder; AG, Acute gastritis; ALS, Amyotrophic lateral sclerosis; AP, Acute pancreatitis; ASD, Autistic spectrum disorders; BC, Breast cancer; BD, Bipolar disorder; CAD, Coronary artery disease; CHD, Coronary heart disease; CKD, Chronic kidney disease; CNS, Central nervous system; CP, Chronic pancreatitis; CRC, Colorectal cancer; CVS, Cardiovascular and cerebrovascular diseases; DKD, Diabetic kidney disease; DS, Digestive system; EC, Endometrial cancer; ED, Erectile dysfunction; EOC, Epithelial ovarian cancer; ES, Endocrinium diseases; GAD, Generalized anxiety disorder; GBM, Glioblastoma multiforme; GBS, Guillain-barré syndrome; GRED, Gastroesophageal reflux disease; GS, Gastric ulcer; GU, Genitourinary diseases; HBV, Hepatitis B virus; HF, Heart failure; HTN, Hypertension; IBS, Irritable bowel syndrome; IDs, Infectious diseases; MDD, Major depressive disorder; MG, Myasthenia gravis; MI, Myocardial infarction; MS, Multiple sclerosis; NAFLD, Nonalcoholic fatty liver disease; NMOSD, Neuromyelitis optica spectrum disorders; OCD, Obsessive compulsive disorder; PAD, Peripheral artery disease; PCOS, Polycystic ovary syndrome; PD, Parkinson's disease; PE, Pulmonary embolism; RA, Rheumatoid arthritis; NRVD, Non-rheumatic valve diseases; RHD, Rheumatic heart disease; SCZ, Schizophrenia; SLE, Systemic lupus erythematosus; T2DM, Type 2 diabetes mellitus; TNFR1, Tumor necrosis factor receptor 1; TNFSF14, Tumor necrosis factor ligand superfamily member 14; TS, Tourette’s syndrome; Tu, Tumour; UC, Ulcerative colitis; VHD, Valvular heart disease; VTE, Venous thrombus embolism.

eTable 4 Preferred Reporting Items for Systematic Reviews and Meta-Analyses

| **Section and Topic** | **Item #** | **Checklist item** | **Location where item is reported** |
| --- | --- | --- | --- |
| **TITLE** | | |  |
| Title | 1 | Identify the report as a systematic review. | Title |
| **ABSTRACT** | | |  |
| Abstract | 2 | See the PRISMA 2020 for Abstracts checklist. | The abstract (Page 2) |
| **INTRODUCTION** | | |  |
| Rationale | 3 | Describe the rationale for the review in the context of existing knowledge. | Introduction (Page 4) |
| Objectives | 4 | Provide an explicit statement of the objective(s) or question(s) the review addresses. | Introduction (Page 4-5) |
| **METHODS** | | |  |
| Eligibility criteria | 5 | Specify the inclusion and exclusion criteria for the review and how studies were grouped for the syntheses. | Methods sub-section: “Literature Search and Inclusion Criteria” (Page 5-6) |
| Information sources | 6 | Specify all databases, registers, websites, organisations, reference lists and other sources searched or consulted to identify studies. Specify the date when each source was last searched or consulted. | Methods sub-section: “Literature Search and Inclusion Criteria” (Page 5-6) |
| Search strategy | 7 | Present the full search strategies for all databases, registers and websites, including any filters and limits used. | Methods sub-section: “Literature Search and Inclusion Criteria” (Page5- 6) |
| Selection process | 8 | Specify the methods used to decide whether a study met the inclusion criteria of the review, including how many reviewers screened each record and each report retrieved, whether they worked independently, and if applicable, details of automation tools used in the process. | Methods sub-section: “Literature Search and Inclusion Criteria”(Page5- 6) |
| Data collection process | 9 | Specify the methods used to collect data from reports, including how many reviewers collected data from each report, whether they worked independently, any processes for obtaining or confirming data from study investigators, and if applicable, details of automation tools used in the process. | Methods sub-section: “Literature Search and Inclusion Criteria” (Page5- 6) |
| Data items | 10a | List and define all outcomes for which data were sought. Specify whether all results that were compatible with each outcome domain in each study were sought (e.g. for all measures, time points, analyses), and if not, the methods used to decide which results to collect. | Methods sub-section: “Data Extraction and Quality Assessment” (Page6) |
|  | 10b | List and define all other variables for which data were sought (e.g. participant and intervention characteristics, funding sources). Describe any assumptions made about any missing or unclear information. | Methods sub-sections: “Data Extraction and Quality Assessment” and “Data Analysis” . No assumptions mad about missing /unclear information. (Page 6-7) |
| Study risk of bias assessment | 11 | Specify the methods used to assess risk of bias in the included studies, including details of the tool(s) used, how many reviewers assessed each study and whether they worked independently, and if applicable, details of automation tools used in the process. | Methods sub-section: “Data Analysis”(Page 6-7) |
| Effect measures | 12 | Specify for each outcome the effect measure(s) (e.g. risk ratio, mean difference) used in the synthesis or presentation of results. | Methods sub-section: “Data Analysis” (Page 6-7) |
| Synthesis methods | 13a | Describe the processes used to decide which studies were eligible for each synthesis (e.g. tabulating the study intervention characteristics and comparing against the planned groups for each synthesis (item #5)). | Methods sub-section: “Data Analysis” (Page 6-7) and Supplementary material etable 1 |
|  | 13b | Describe any methods required to prepare the data for presentation or synthesis, such as handling of missing summary statistics, or data conversions. | Methods sub-section: “Data Analysis” . (Page 6-7) No missing summary statistics. |
|  | 13c | Describe any methods used to tabulate or visually display results of individual studies and syntheses. | Methods sub-section: “Data Analysis” . (Page 6-7) |
|  | 13d | Describe any methods used to synthesize results and provide a rationale for the choice(s). If meta-analysis was performed, describe the model(s), method(s) to identify the presence and extent of statistical heterogeneity, and software package(s) used. | Methods sub-section: “Data Analysis” (Page 6-7) |
|  | 13e | Describe any methods used to explore possible causes of heterogeneity among study results (e.g. subgroup analysis, meta-regression). | Methods sub-section: “Data Analysis” (Page 6-7) |
|  | 13f | Describe any sensitivity analyses conducted to assess robustness of the synthesized results. | Methods sub- section: “Data Analysis” (Page 6-7) |
| Reporting bias assessment | 14 | Describe any methods used to assess risk of bias due to missing results in a synthesis (arising from reporting biases). | Methods sub- section: “Data Analysis” (Page 6-7) |
| Certainty assessment | 15 | Describe any methods used to assess certainty (or confidence) in the body of evidence for an outcome. | Methods sub- section: “Data Analysis” (Page 6-7) |
| **RESULTS** | | |  |
| Study selection | 16a | Describe the results of the search and selection process, from the number of records identified in the search to the number of studies included in the review, ideally using a flow diagram. | Flowchart in Figure 1 |
|  | 16b | Cite studies that might appear to meet the inclusion criteria, but which were excluded, and explain why they were excluded. | Results section “Literature search, study selection” (Page 7)and Flowchart in Figure 1 |
| Study characteristics | 17 | Cite each included study and present its characteristics. | Supplementary Material eTable1 |
| Risk of bias in studies | 18 | Present assessments of risk of bias for each included study. | Supplementary Material eTable2 |
| Results of individual studies | 19 | For all outcomes, present, for each study: (a) summary statistics for each group (where appropriate) and (b) an effect estimate and its precision (e.g. confidence/credible interval), ideally using structured tables or plots. | Supplementary Material eTable3 |
| Results of syntheses | 20a | For each synthesis, briefly summarise the characteristics and risk of bias among contributing studies. | Results section “Study Description” (Page 7) |
|  | 20b | Present results of all statistical syntheses conducted. If meta-analysis was done, present for each the summary estimate and its precision (e.g. confidence/credible interval) and measures of statistical heterogeneity. If comparing groups, describe the direction of the effect. | Results section “Meta-  Analysis Results” , (Page 7-8) Supplementary Material eTable 3 |
|  | 20c | Present results of all investigations of possible causes of heterogeneity among study results. | Results section “Meta-Analysis Results”, (Page 7-8) Supplementary Material eTable 3 |
|  | 20d | Present results of all sensitivity analyses conducted to assess the robustness of the synthesized results. | Results section “Meta-Analysis Results”, (Page 7-8)Supplementary Material eTable 3 and Table 2 |
| Reporting biases | 21 | Present assessments of risk of bias due to missing results (arising from reporting biases) for each synthesis assessed. | NA |
| Certainty of evidence | 22 | Present assessments of certainty (or confidence) in the body of evidence for each outcome assessed. | Supplementary Material eTable 3 and Table 1 |
| **DISCUSSION** | | |  |
| Discussion | 23a | Provide a general interpretation of the results in the context of other evidence. | Discussion section (Page 10-12) |
|  | 23b | Discuss any limitations of the evidence included in the review. | Discussion section (Page 10-12) |
|  | 23c | Discuss any limitations of the review processes used. | Discussion section (Page 10-12) |
|  | 23d | Discuss implications of the results for practice, policy, and future research. | Discussion section (Page 10-12) |
| **OTHER INFORMATION** | | |  |
| Registration and protocol | 24a | Provide registration information for the review, including register name and registration number, or state that the review was not registered. | Abstract and methods (Page 2, 5-7) |
|  | 24b | Indicate where the review protocol can be accessed, or state that a protocol was not prepared. | NA |
|  | 24c | Describe and explain any amendments to information provided at registration or in the protocol. | NA |
| Support | 25 | Describe sources of financial or non-financial support for the review, and the role of the funders or sponsors in the review. | Abstract and “Acknowledgments” subsection (Page 2, 13) |
| Competing interests | 26 | Declare any competing interests of review authors. | “Declaration of interests” sub-section (Page 13) |
| Availability of data, code and other materials | 27 | Report which of the following are publicly available and where they can be found: template data collection forms; data extracted from included studies; data used for all analyses; analytic code; any other materials used in the review. | Extraction sheet within linked protocol |
